# Fronto-central resting-state 15-29Hz transient beta events change with therapeutic transcranial magnetic stimulation for posttraumatic stress disorder and major depressive disorder

**DOI:** 10.1101/2023.03.11.23286902

**Authors:** Alexander T. Morris, Simona Temereanca, Amin Zandvakili, Ryan Thorpe, Danielle D. Sliva, Benjamin D. Greenberg, Linda L. Carpenter, Noah S. Philip, Stephanie R. Jones

**Affiliations:** VA RR&D Center for Neurorestoration and Neurotechnology, VA Providence, Providence, RI; Department of Psychiatry and Human Behavior, Alpert Medical School of Brown University, Providence, RI; COBRE Center for Neuromodulation, Butler Hospital, Providence, RI; Department of Neuroscience, Brown University, Providence, RI; Carney Institute for Brain Science, Brown University, Providence, RI

## Abstract

Repetitive transcranial magnetic stimulation (rTMS) is an established treatment for major depressive disorder (MDD) and shows promise for posttraumatic stress disorder (PTSD), yet effectiveness varies. Electroencephalography (EEG) can identify rTMS-associated brain changes. EEG oscillations are often examined using averaging approaches that mask finer time-scale dynamics. Recent advances show some brain oscillations emerge as transient increases in power, a phenomenon termed “Spectral Events,” and that event characteristics correspond with cognitive functions. We applied Spectral Event analyses to identify potential EEG biomarkers of effective rTMS treatment. Resting 8-electrode EEG was collected from 23 patients with MDD and PTSD before and after 5Hz rTMS targeting the left dorsolateral prefrontal cortex. Using an open-source toolbox (https://github.com/jonescompneurolab/SpectralEvents), we quantified event features and tested for treatment associated changes. Spectral Events in delta/theta (1-6 Hz), alpha (7-14 Hz), and beta (15-29 Hz) bands occurred in all patients. rTMS-induced improvement in comorbid MDD PTSD were associated with pre-to post-treatment changes in fronto-central electrode beta event features, including frontal beta event frequency spans and durations, and central beta event maxima power. Furthermore, frontal pre-treatment beta event duration correlated negatively with MDD symptom improvement. Beta events may provide new biomarkers of clinical response and advance the understanding of rTMS.

## INTRODUCTION

Posttraumatic stress disorder (PTSD) and major depressive disorder (MDD) are prevalent conditions that substantially degrade functioning and quality of life. MDD and PTSD diagnoses are extensive in the United States, with diagnoses of either reaching over 10% in a large sample representative of the general population and over 28% in a large Veteran population [1,2]. These conditions are highly comorbid; up to 50% of patients with PTSD are also diagnosed with MDD [3,4]. While each disorder can be difficult to treat individually [5,6], patients with comorbid depression and anxiety symptoms have poorer treatment outcomes [7,8] and standard depression treatments are less effective when PTSD is present [9,10].

Neuromodulation using repetitive transcranial magnetic stimulation (rTMS) has emerged as an efficacious treatment for pharmacoresistant MDD [11] and MDD comorbid with PTSD [12]. However, therapeutic effects of rTMS are variable [13]. Understanding how rTMS affects brain circuits and how individual variation in oscillatory activity influences the therapeutic mechanisms of rTMS may improve treatment outcomes for patients with complex phenotypes or comorbid syndromes.

Electroencephalography (EEG) provides a powerful means to evaluate the effect of rTMS treatments on fast time-scale brain dynamics [14,15]. Data analysis techniques applied to evaluate the impact of rTMS on EEG signals commonly rely on quantifying changes in the spectral domain, such as changes in regional oscillatory power and/or coherence among brain areas. Candidate EEG biomarkers to assess the effects of rTMS for depression have been proposed across several frequency bands (typically <40Hz), and for a range of spectral features, including averaged local band power [16,17,18], event-related power and coherence [19,20], and for varied measures of time-domain signal complexity [21]. Recent studies applying machine learning showed that rTMS-induced changes in resting-state EEG coherences were predictive of clinical response in comorbid PTSD/MDD and able to distinguish active, sham, pre-treatment, and post-treatment groups [22,23].

Spectral EEG analyses, including all those referenced above, typically rely on Fourier methods that assume the signal can be well represented by continuous sinusoidal oscillations that remain invariant and are averaged across time. Yet, challenges remain related to the replicability and application of previously observed findings [24]. In recent years, there has been a paradigm shift in the application of these methods, as many studies have now shown that, in unaveraged data, brain oscillations often occur as transient increases in high spectral power, a phenomenon termed oscillatory “bursts” or “events” [25,26]. Quantification of transient changes in spectral activity requires new methods that consider temporal characteristics of spectral activity such as event rates, amplitudes, durations, or frequency spans [27]. Such “Spectral Event” methods have recently been applied in an increasing body of EEG (and/or MEG) studies and are leading to new insights on brain dynamics of sensory information processing [27,28], motor action [29,30,31], working memory [32,33], and neuropathology [34,35,36,37,38]. However, to our knowledge, such Spectral Event analyses have not yet been applied to quantify the impact of rTMS on EEG-measured brain dynamics and may provide a new pathway to defining EEG biomarkers of rTMS treatment efficacy.

We applied Spectral Event analysis methods developed by our group (https://github.com/jonescompneurolab/SpectralEvents; [27]) to resting-state EEG datasets from adults with comorbid MDD and PTSD who received 5Hz rTMS over the left dorsolateral prefrontal cortex (DLPFC) in an open-label trial. We hypothesized that transient high-power spectral “Events” would be present in the resting-state theta/delta (1-6 Hz), alpha (7–14 Hz) and beta (15–29 Hz) frequency bands and that clinical improvement after rTMS treatment would be associated with one or more event features.

## RESULTS

Patients (56.5% male, 91.3% Caucasian, mean age 52.5±9.5) received up to 40 sessions of 5Hz TMS to left DLPFC to treat comorbid MDD and PTSD. Resting-state eyes-closed EEG data were recorded from eight electrodes before and after treatment (see Fig. 1).

**Figure 1:**
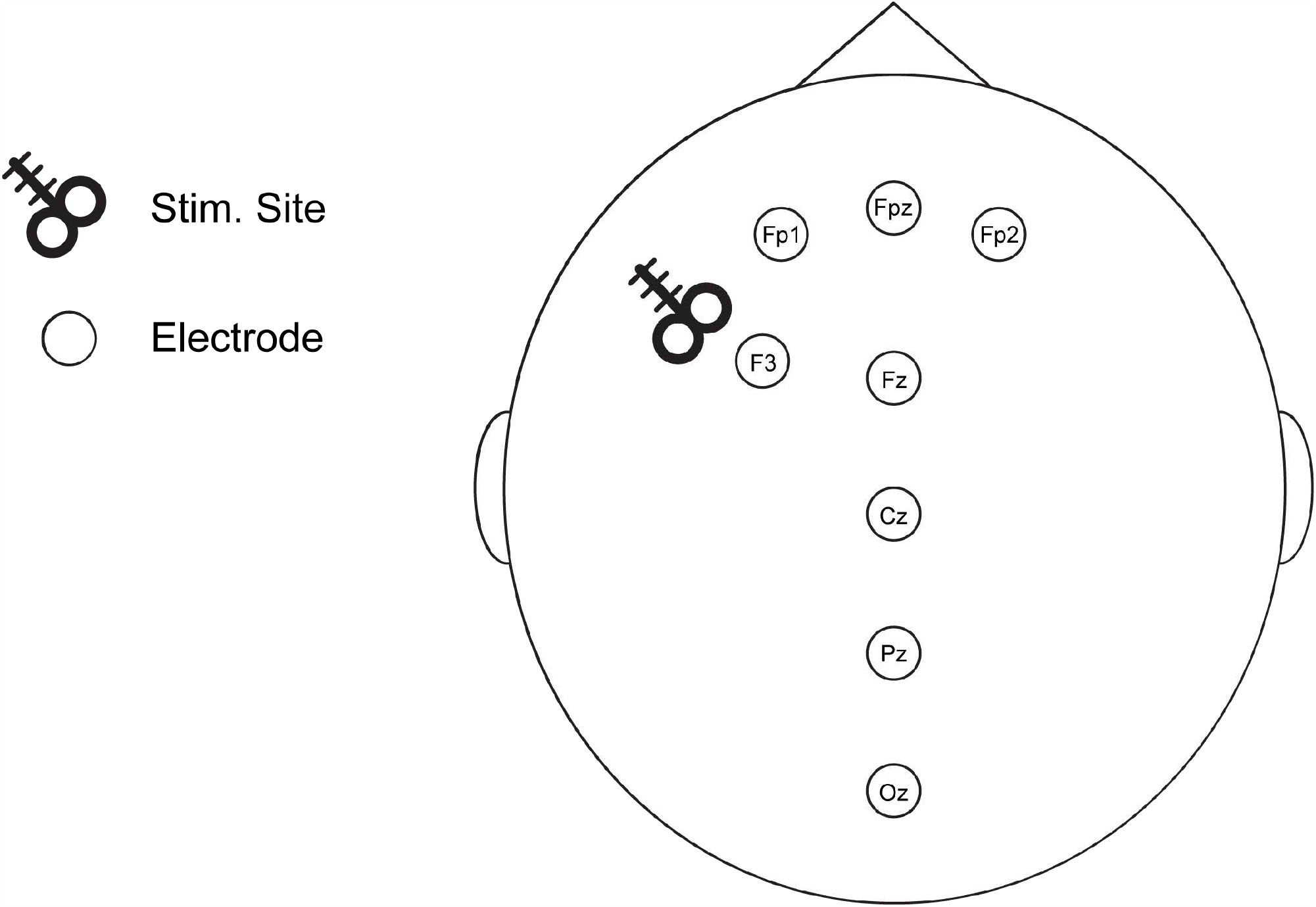
EEG electrode placement and TMS stimulation site. Locations of 8 electrodes, including the stimulation site targeting the left DLPFC (over F3).

### PTSD and MDD Symptoms Improved after rTMS

The sample showed a mean 39.0% PCL-5 score reduction and a 39.3% IDS-SR score reduction after rTMS. Figure 2 shows clinical improvement for each patient. The course of rTMS treatment led to a clinical response (PCL-5 raw score reduction >10 points) in 18/23 patients for PTSD symptoms, and clinical response (IDS-SR percent score reduction ≥50%) in 12/23 patients for MDD symptoms. PCL-5 score change was highly correlated with IDS-SR percent change across patients (r = 0.874, p < 0.0001, data not shown).

**Figure 2:**
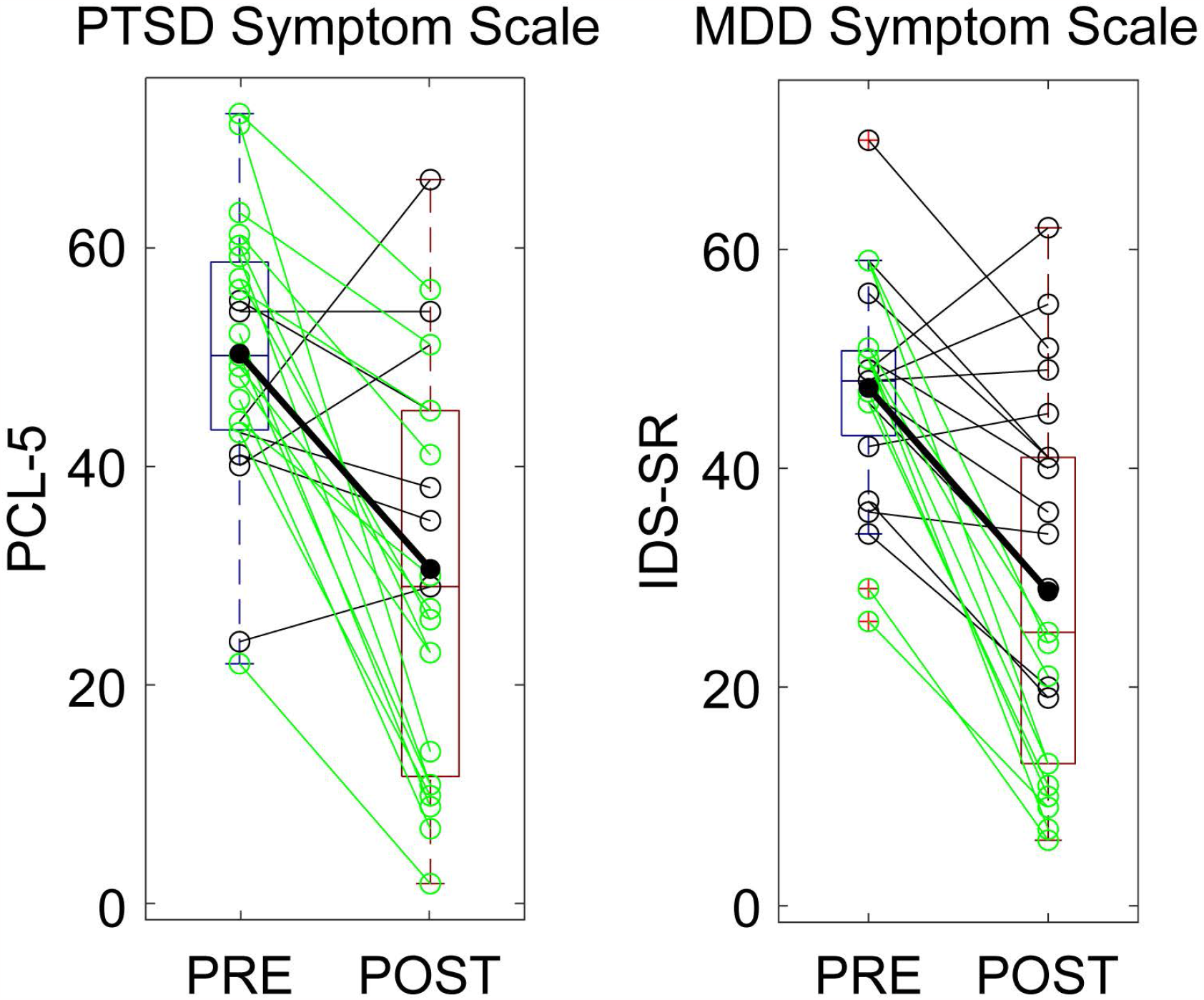
Clinical score change pre-(blue) to post-treatment (red) for all patients. Scores reported for tahe PTSD symptom scale (PCL-5, left) and the MDD symptom scale (IDS-SR, right) for patients with a clinical response (green) and without (black) as well as the group average (bold).

### EEG Power Spectral Density (PSD) Revealed Peaks in Delta/Theta, Alpha, and Beta Frequency Bands Pre- and Post-Treatment

To define frequency ranges for our Spectral Event analyses in a principled manner, we first performed a PSD analysis of the EEG signals to determine high-power frequency ranges. Figure 3 presents power grand averages and standard error of the mean bars across all patients and EEG recording sessions (i.e., pre-and post-treatment). For each EEG sensor, peaks in activity consistently appear in delta/theta (1-6 Hz), alpha (7-14 Hz), and beta range (15-29 Hz). As such, these bands defined the frequency boundaries for all further analyses.

**Figure 3:**
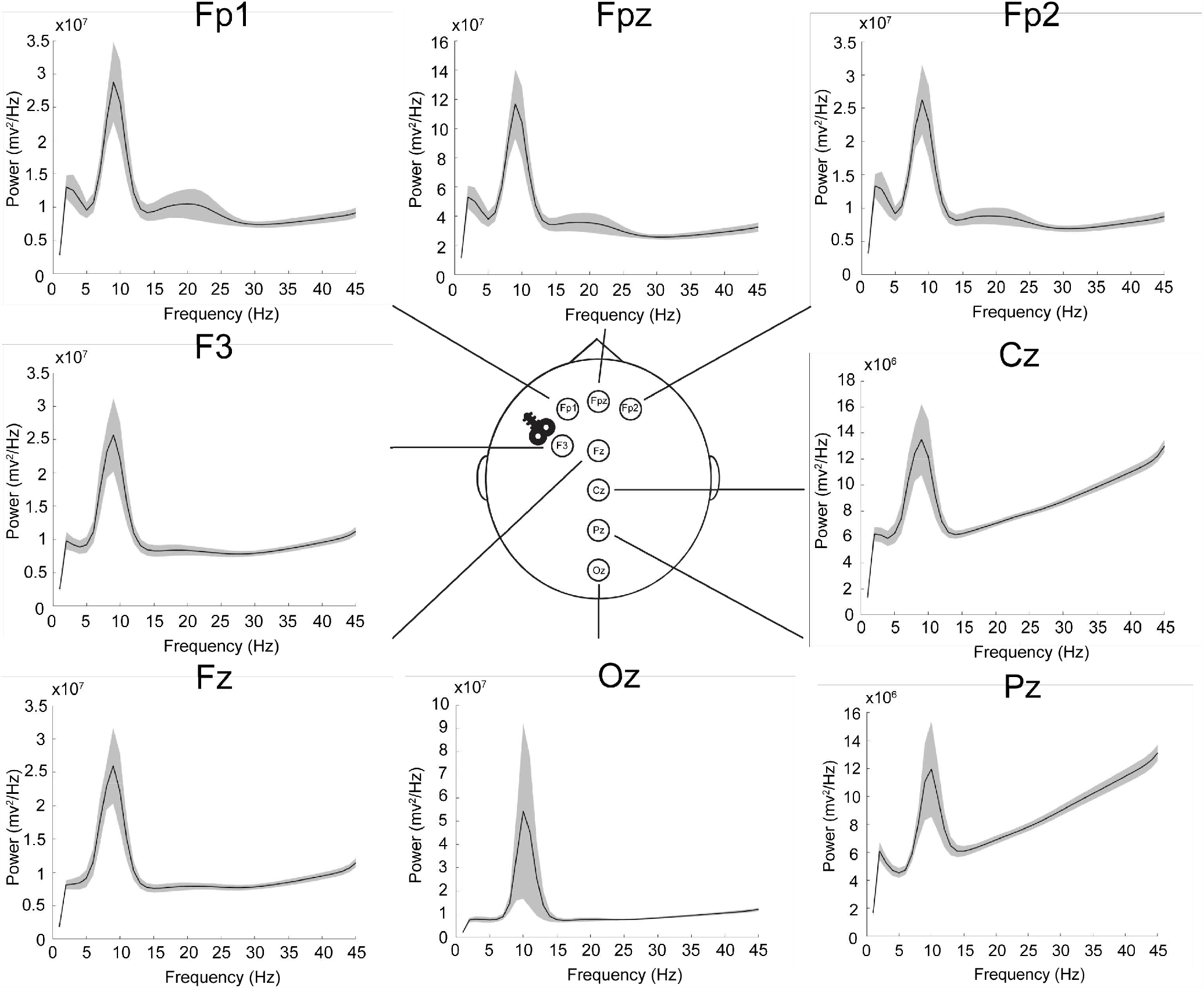
Power spectral density plots averaged across patients and EEG recordizng sessions for each electrode. Mean (solid line) and one standard error from the mean (transparent volumes) reported.

### Averaged PSD was Unchanged Post-Treatment

To compare effects of rTMS treatment on average power, we averaged the signal power in each identified band of interest (BOI). For each band, we performed a repeated measures two-way ANOVA with factors of time (pre- and post-treatment) and electrode (eight EEG electrodes) across all 23 patients. There were no main effects of time or interaction (F(7,22) < 3.664, p > 0.05 for all variables), indicating similar average power in each BOI and electrode pre- and post-treatment.

### Transient Delta/Theta, Beta and Alpha Events Were Detected and Quantified in Pre- and Post-Treatment Resting State EEG Data

Spectral Event analysis of unaveraged 2-second epochs of resting state EEG signals revealed transient high-power events in each BOI (see examples in Fig. 4a, and Supplementary Fig. S2). The average Time-Frequency Response (TFR) across epochs (Fig. 4a upper left) showed continuous bands of high power in delta/theta (1-6 Hz), alpha (7-14 Hz), and beta (15-29 Hz) activity due to the accumulation of events that occurs during averaging. Example unaveraged epochs revealed transient increases in high power activity, i.e., Spectral Events, within each BOI. We quantified the following event features: number of events per 2-sec epoch, event maxima power, event duration, and event frequency span. Histograms for beta event Features for individual patients are shown in Fig. 4b and quantified for each BOI in Fig S2 and Table S1. Across all patients, in the delta/theta range there were on average 0.69±0.03 events per 2-sec epoch, with 2.41±0.38×10^7^ mV^2^/Hz maxima power, 884.60±25.25 ms duration, and 3.09±0.16 Hz frequency span. In the alpha range, there were 1.28±0.05 events per 2-sec epoch, with 3.38±0.49×10^7^ mV^2^/Hz maxima power, 295.36±7.72 ms duration, and 5.49±0.24 Hz frequency span. In the beta range, there were 2.35±0.07 events per 2-sec epoch, with 2.23±0.16×10^7^ mV^2^/Hz maxima power, 136.87±1.54 ms duration, and 9.82±0.18 Hz frequency span. Beta event values were consistent with prior analysis of resting state data [36].

**Figure 4:**
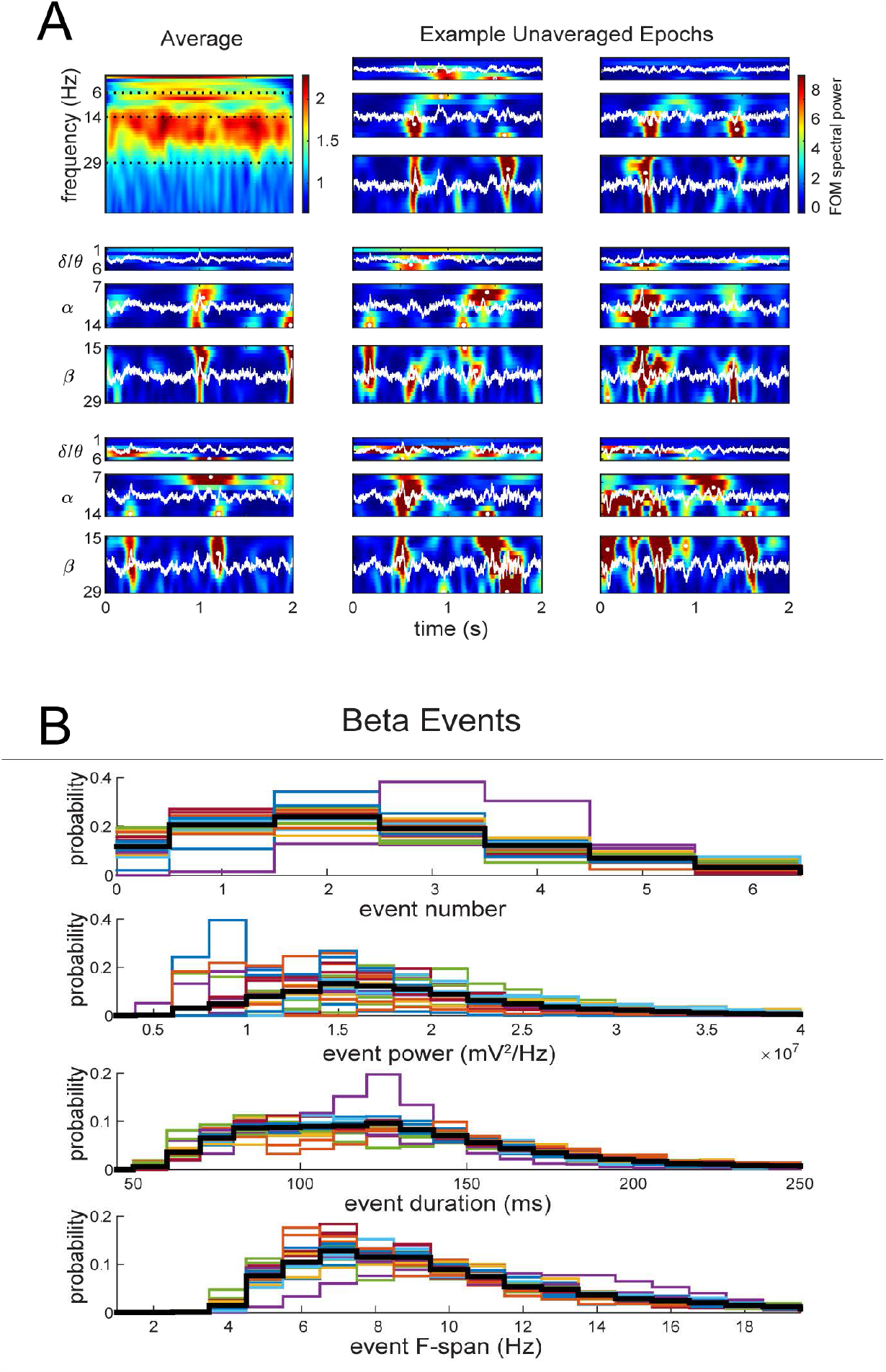
Low-frequency rhythms emerge as quantifiable transient high-power events (white dots) from non-averaged time-series waveform (white line) converted into TFRs. (a) Spectrogram showing average Fpz electrode power (normalized by the median value of each frequency) over frequency and time across all pre-treatment epochs from an example patient (top left), and time-frequency spectrograms of eight randomly chosen individual epochs for each BOI. Color bar denotes heat map of power values as factors of the median (FOM) spectral power. (b) Histograms showing the distribution of all beta event feature measurements from the Fpz electrode for individual patients (colored lines) and the average (black line). Event number is counted over 2 second epochs. See Supplementary Info for further Spectral Event feature quantification across frequency bands (Fig. S.2) and across all electrodes (Table S.1).

We next examined if rTMS impacted event features, regardless of clinical outcome, by pooling the data from all patients and comparing event features before and after rTMS treatments. When averaging across all patients, no significant differences between pre- and post-treatment measurements were found for any event feature. See Supplementary Table S2 for p-values and percent change values from all bands and electrodes, as well as Supplementary Table S1 for average totals of each event feature from each electrode.

### Changes in Fronto-Central Electrode Beta Event Features Correlated with Clinical Improvement

While there were no significant pre-to post-treatment differences in event features when averaging the data across all patients, we hypothesized that pre-to post-treatment changes in event features may depend on clinical outcomes. We found that changes in frontal and central electrode beta event features significantly correlated with clinical improvements (Fig. 5). Significant correlations were not found in any other BOI (Table S3).

**Figure 5:**
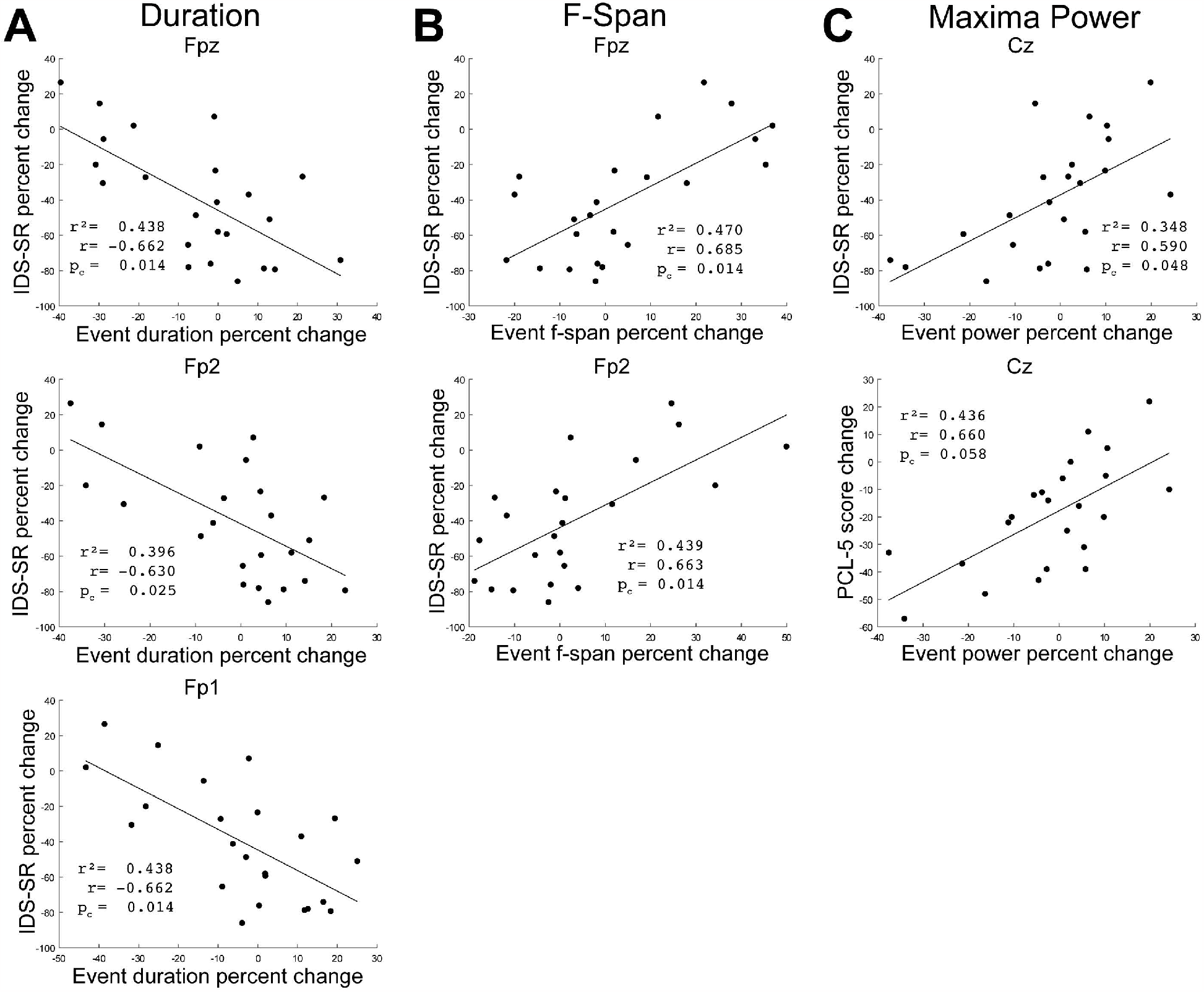
Correlation between change in clinical score and beta event features pre-to post-treatment for each patient (•). (a) Percent change in beta event duration vs. IDS-SR percent change for the Fpz, Fp2, and Fp1 electrodes. (b) Percent change in beta event frequency span vs. IDS-SR percent change for the Fpz and Fp2 electrodes. (c) Percent change in beta maxima power vs. IDS-SR percent change and PCL-5 score change for the Cz electrode. Pearson’s correlation coefficient r, r^2^, and BH-adjusted p-values reported.

Patients that reported post-treatment improvement in MDD symptoms (i.e., decreased IDS-SR scores) exhibited increased beta event durations in frontal electrodes, while those with worse outcomes exhibited decreased durations (significant correlations in pre-to post-changes: Fpz: r = -0.662, p_c_ = 0.014; Fp2: r = -0.630, p_c_ = 0.025; Fp1: r = -0.662, p_c_ = 0.014, where p_c_ is the Benjamini-Hochberg adjusted p-value corrected for multiple comparisons across the correction space of event features, electrodes, and BOI) (Fig. 5a). Moreover, MDD symptom improvement was associated with decreased post-treatment frontal beta event frequency span (Fig 5b; Fpz: r = 0.685; p_c_ = 0.014, Fp2: r = 0.663, p_c_ =0.014), and decreased central electrode beta event maxima power (Fig 5c top; Cz: r = 0.590; p_c_ = 0.048, with an additional trend toward decreased power in neighboring electrode Fz: r=0.571, p_c_ = 0.060, see Table S3).

Patients reporting improvements in PTSD symptoms (decreased PCL-5 scores) similarly exhibited a trend towards decreased central beta event maxima power (Fig 5c bottom; Cz: r = 0.660, p_c_ = 0.058), though this result was not statistically significant with our False Discovery Rate set at 0.05.

See Supplementary Figs. S3 and S4 for further illustration of our findings.

### Pre-Treatment Beta Event Duration in Frontal Electrodes Correlates with Clinical Response to rTMS

To determine if event features at baseline were associated with post-treatment symptom improvements from rTMS, we calculated the correlation between pre-treatment event feature measurements and clinical score change separately for each BOI and electrode. Patients that reported post-treatment IDS-SR symptom improvement (decreased scores) had shorter pre-treatment beta event durations, while those with worse outcomes had longer pre-treatment durations (Fig. 6, significant correlations Fpz: r = 0.683, p_c_ = 0.023; Fp2: r = 0.659, p_c_ = 0.023). Significant effects were not found for the PCL-5 or in any other BOI (Table S4).

**Figure 6:**
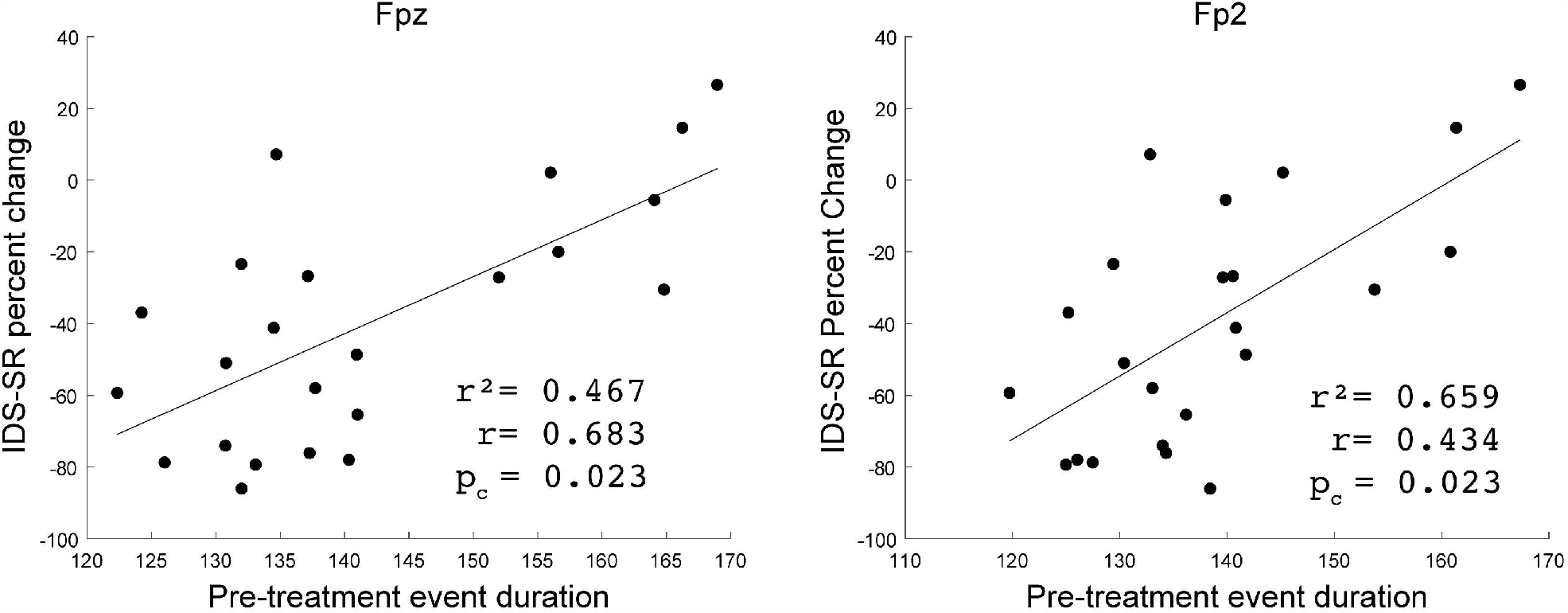
Correlation between IDS-SR percent change and pre-treatment beta event duration from the Fpz and Fp2 electrodes for each patient (•). Pearson’s correlation coefficient r, r^2^, and BH-adjusted p-values reported.

## DISCUSSION

This study implemented EEG Spectral Event methods to quantify features of transient high-power activity in unaveraged EEG data and to study oscillatory changes occurring in relation to clinical efficacy of rTMS in patients with comorbid MDD and PTSD [12]. Using a sparse 8-electrode montage, we found resting-state EEG oscillations exhibited transient high-power events in the delta/theta, alpha, and beta ranges before and after treatment (Fig. 4). While transient events have been detected in resting-state data [28,36,37,42,43,44], this study is the first to identify and quantify them in a population with MDD and PTSD.

To our knowledge, prior studies have not examined the effects of TMS on Spectral Event features. We found that clinical improvement in MDD and PTSD was associated with decreased frequency spans and increased durations of frontal electrode beta events, and with decreased maxima event power in central electrodes (Fig. 5). Further, shorter frontal electrode beta event durations at the pre-treatment baseline were correlated with greater post-treatment improvement in depression symptoms (Fig. 6). Taken together, the results surrounding beta event durations suggest that patients starting with shorter beta event durations are most likely to exhibit a pre-to post-treatment duration increase reflective of symptom improvement. These findings merit replication in other rTMS samples with MDD and PTSD, as well as exploration in samples with singular diagnosis of either MDD or PTSD.

By moving beyond trial averaging, transient beta events may provide a new potential biomarker for effective rTMS treatment. Spectral Event analyses can increase statistical power that is lost with averaging across time and/or trials and may help overcome issues of stability and reproducibility, which are a major limiting factor in establishing EEG as a reliable measure of therapeutic rTMS treatments [24,45]. Consistent with prior studies [20,46,47,48], including those using the dataset studied here [22], we did not find differences in averaged EEG power with rTMS treatment for PTSD or MDD. The Spectral Event analysis provided a finer-grained investigation that revealed changes in beta band activity, not previously reported.

More advanced methods that go beyond power calculations to quantify coordination of oscillations across brain regions have shown more promise for rTMS response prediction ([49],[50],[51], reviewed in [52]). We have previously applied machine learning classification to show that across-electrode spectral coherence of EEG resting state oscillations in the alpha, beta, theta and delta bands are predictive of symptom improvements in the 5Hz TMS patient group studied here [22]. In the beta band, coherence between frontal and midline electrodes played the strongest role in classification performance (see also, [49], [51]). The current beta event findings suggest that the previously reported coherence may be related to the across area coordination of transient beta events, as has been observed in other resting state studies with methods that are beyond the scope of the current study [31].

A classic interpretation of therapeutic rTMS indicates that its mechanisms involve modulation in the plasticity of brain circuits and networks [53,54]. How this plasticity takes shape and leads to symptom improvement is unknown. The observed beta event feature changes with therapeutic rTMS may provide unique insight into this important open question.

The sparse electrode array used in this study limits conclusions that can be made about the precise anatomical location of the cortical circuits underlying our observations. However, the beta event frequency span and duration results observed in frontal electrodes are distinct from the maxima event power results found in central electrodes, suggesting they emerge from anatomically distinct underlying networks. Moreover, the wavelet methods applied to calculate the time-frequency representations of the data (e.g. Fig. 4a) impose a trade-off between time and frequency resolution, and hence a potential coupling between longer durations and shorter frequency spans, supporting the notion that the frontal electrode results are associated with the same underlying neural process.

Beta rhythms in frontocentral executive control regions have been implicated in higher order cognitive processes, e.g. cognitive control and working memory [32,33,55], with direct links to symptom severity in MDD and PTSD [56,57,58,59]. Averaged beta power increases in executive control regions are often associated with the “inhibition” of distracting information [30,57,60] or emotions [59], and with the suppression of motor actions [29,30,61,62,63,64,65,66,67].

Recent findings directly link beta-associated inhibitory influences to changes in beta event (i.e., burst) features. Several stop-signal task studies have shown that beta event rate in frontal cortex increases with successful cancellation of action, and that the timing of beta events correlates with reaction times, such that earlier events lead to faster stop times [62,63,64,65,66,67]. Transient beta events in frontal regions have also been associated with working memory and decision making, where it has been suggested that they support the formation of neural ensembles needed to meet current task demands [32,33].

All of these prior studies focus on beta event timing and rate. To our knowledge, none have directly related changes in beta event duration, frequency span, or maximum event power with cognition or inhibitory control. Our results suggest the TMS-induced changes in these beta event features may also be reflective of the suppression of distracting thoughts, emotions and/or memories, which lead to improved MDD and PTSD symptoms.

The observed rTMS induced beta event feature changes may also provide novel insight into the biophysical neural mechanisms through which beta events regulate inhibitory control. A recent computational modeling-derived theory by our group on the biophysical cellular and circuit-level neural mechanisms generating beta events provides a potential interpretation of the mechanisms through which beta event related “inhibition” of thoughts, emotions, and/or memories may occur [68,69,70]. In brief, we have observed similarities in beta event features in sensory, frontal, and motor cortices [60,68,71]. Using an integrated approach that combined neural modeling with human, non-human primate, and mouse recordings, we developed a theory on the mechanisms of beta event generation. This theory suggested beta events emerge from the integration of layer-specific thalamocortical excitation of the neocortical column, where a strong ∼50ms (i.e., beta period) burst of supragranular excitation from thalamus to the distal dendrites of neocortical pyramidal neurons was the most prominent defining mechanism of beta event generation [68,71]. Follow-up studies further predicted that the supragranular excitation also recruits inhibitory neurons, whose activity suppresses the relay of incoming sensory information leading to decreased sensory perception [27, 70]. How these mechanisms relate to the observed TMS-induced changes in beta event frequency span, duration and maxima event power requires further investigation beyond the scope of the current study. However, based on our prior beta event generation theory, we speculate that beta-associated inhibitory neuron recruitment may also underlie the suppression of negative thoughts and emotions in MDD and PTSD. Moreover, rTMS may induce changes in thalamocortical connectivity plasticity, mediated in part by its impact on inhibitory neurons [72,73].

In conclusion, our findings serve as a critical first step in defining novel EEG Spectral Event feature biomarkers of therapeutic rTMS for MDD and PTSD. These results are meant to be fundamental in nature providing new insights into the mechanisms of action of rTMS. At present they cannot guide clinical practice. However, they lay the foundation for further replication studies and development of a deeper understanding of the circuit-level mechanisms underlying how rTMS induces changes in Spectral Event features that may ultimately be a useful clinical biomarker to guide treatment.

## METHODS

### TMS Paradigms and Patient Populations

We analyzed data from 23 patients from a previously published clinical study conducted at the VA Medical Center and Butler Hospital in Providence, RI, USA. The full study cohort included thirty-five patients diagnosed with moderate to severe comorbid MDD+PTSD. Patients received a course of once-daily (weekdays) unblinded 5Hz rTMS stimulation sessions, delivered to the left DLPFC, for up to 40 sessions. Daily treatments each included 3000-4000 pulses, with treatment intensity delivered at 120% of the motor threshold on a NeuroStar device system (Neuronetics Inc., Malvern PA). The stimulation protocol was comprised of 5s trains and 14s inter-train intervals (NCT02273063; for full details see [12]). EEG data was acquired at baseline (pre-treatment) and after the last TMS session (post-treatment). The stimulation site (F3, Fig. 1) over the left DLPFC was determined using the Beam/F3 method [39].

Symptom severity for MDD and PTSD was evaluated using validated self-report measures: the Inventory of Depressive Symptoms-Self Report (IDS-SR) [40] for MDD and the PTSD Checklist for DSM-5 (PCL-5) [41]. For each report, a decrease in score indicates an improvement in symptoms. All patients met clinical and safety criteria for rTMS. Concurrent psychotropic medications and ongoing psychotherapy were continued at stable regimens during participation. Written informed consent was obtained for all study procedures from all participants. All procedures were performed in accordance with current guidelines and regulations and were approved by the VA Providence and Butler Institutional Review Boards.

### EEG acquisition

Details of the EEG acquisition was previously reported [22]. Briefly, resting-state eyes-closed EEG was recorded while patients sat quietly in a sound-attenuated room. Patients were asked to keep eyes open for one minute, closed for 10–12 minutes, and open again for another minute. Only eyes-closed data were analyzed. An 8-channel electrode cap and EEG device (ENOBIO8, Neuroelectrics, Cambridge, MA, USA) were employed to record data from dry electrodes placed over FP1, FP2, FPz, F3, Fz, Cz, Pz, and Oz (Fig. 1). EEG acquisition used a high-pass (0.5 Hz) and low-pass (50 Hz) filter, with sampling at 500 Hz and 24-bit precision digitization.

### EEG analysis

EEG preprocessing was performed using a similar approach as in [22] with custom MATLAB code (v2019a; Mathworks, Natick, MA, USA). The data were segmented into 2-second non-overlapping epochs, and epochs containing artifacts such as eye movement, muscle, movement-related, and amplifier drift were removed after manual inspection (masked for clinical outcome status). Only data from individuals with >120 s of usable EEG data (sixty 2-s epochs) from all 8-electrodes and with complete clinical data were used in the analysis, resulting in 23 patients with complete data (66% of the original data set). An equal number of epochs from pre-treatment and post-treatment recording sessions were randomly selected for analysis.

### Power Spectral Density (PSD) analysis

Time-Frequency Responses (TFRs) were calculated using methods as in MATLAB *SpectralEvents* Toolbox (https://github.com/jonescompneurolab/SpectralEvents, findMethod=3) ([37], see also [27]). Each artifact-free 2-sec epoch was convolved with a 7-cycle Morlet wavelet. Power spectral density (1-45 Hz) for each EEG electrode was calculated by averaging the TFR across time for all epochs across both EEG recording sessions and for each patient. We then computed power in theta/delta (1-6 Hz), alpha (7–14 Hz) and beta (15–29 Hz) frequency bands by averaging power within each frequency band of interest (BOI).

### Spectral Event Analysis

Spectral Events were defined using TFRs normalized by the median power value for each frequency value throughout each BOI. Transient high-power “events” were detected and characterized using the Spectral Events Toolbox, defined for each frequency value as local maxima above a 6X median threshold within a BOI (see Supplementary Fig. S1 for additional details). Specifically, for each patient, Spectral Events were found by (1) thresholding normalized TFR in the BOI, using a 6X factor of the median (FOM) threshold computed separately for each individual frequency; and (2) finding all local maxima and discarding those of lesser magnitude in each suprathreshold region, so that only the greatest local maximum in each region survived. In the rare case when more than one local maximum in a region had the same greatest value, their respective event timing, frequency, and boundaries at full-width half-max (see duration and frequency span below) were calculated separately and averaged. This method does not allow for overlapping events to occur in a suprathreshold region and ensures that the presence of within-band, suprathreshold activity in any given trial will be counted as one event. Event *number* was calculated by counting the number of events in the 2-second period of each epoch. Event *power* was calculated as the normalized FOM power value at each event maximum. Event *duration* and *frequency span* were calculated from the boundaries of the region containing power values greater than half the local maxima power, as the full-width-at-half-maximum in the time and frequency domain, respectively. Edge cases in the time and frequency domains were resolved by doubling the half-width of the side that was not cut by the edge.

### Statistical Analysis

Effects of rTMS treatment on average power were assessed for each BOI by performing a two-way repeated measures ANOVA with factors of time (pre- and post-treatment) and electrode (eight EEG electrodes). Ad hoc paired t-tests were performed to evaluate treatment (time) effects for individual electrodes.

For each BOI, paired t-tests were used to assess effects of rTMS treatment on EEG transient event features (event number, power, duration and frequency span) by comparing pre- and post-treatment values for each electrode.

Pearson correlation tests were used to assess the relationships between the pre-to post-treatment percent change in event features and the percent change in MDD and PTSD symptom scores (IDS-SR and PCL-5, respectively), separately for each BOI and electrode. Correlation tests were also used to evaluate (separately for each BOI and electrode) relationships between baseline pre-treatment event features and percent change on symptom scales. All reported p-values were corrected for multiple comparisons across the correction space of event features, electrodes, and BOI (4 × 8 × 3 =96) using the Benjamini–Hochberg (BH) step-up procedure [74] with a False Discovery Rate (Q) set at 0.05. Trending BH-adjusted p-values (a.k.a. the q-values) are reported as 0.05 < p_c_ < 0.08, while statistically significant BH-adjusted p-values are reported as p_c_ < 0.05 (Q = 0.05).

Upon publication, all analysis code will be made publicly available.

## Supporting information

Manuscript

## Data Availability

The datasets generated during and/or analyzed during the current study are not publicly available and may be available from the corresponding author on reasonable request and subject US Department of Veterans Affairs rules on data sharing.

## Acknowledgements

Original trial supported in part by Neuronetics, Inc., through an investigator-initiated grant to LLC (Butler Hospital).

In the last three years, NSP has received clinical trial support (through VA contracts) from Wave Neuro and Neurolief, Inc. LLC has received support (through contracts with Butler Hospital) from Neuronetics, Affect Neuro, Janssen, Neurolief, and Nexstim. ATM, ST, AZ, RT, DDS, BDG, and SRJ have no conflicts of interest.

Effort on this manuscript was supported in part by VA grants I50 RX002864, IK2 CX002115, and I01 CX002088; NIH grants P20 GM130452.

We thank research assistants Dylan Daniels and Elizabeth Kaplan for support in compiling and submitting the manuscript.

The views expressed in this article are those of the authors and do not necessarily reflect the position or policy of the VA or the NIH. The funders had no role in the design of the study, data analysis, or decision to publish.

## REFERENCES

[1] Kessler, R. C., Chiu, W. T., Demler, O., & Walters, E. E. (2005). Prevalence, severity, and comorbidity of 12-month DSM-IV disorders in the national comorbidity survey replication. Archives of General Psychiatry, 62(6), 617. https://doi.org/10.1001/archpsyc.62.6.617

[2] Thomas, J. L., et al. (2010). Prevalence of mental health problems and functional impairment among active component and national guard soldiers 3 and 12 months following combat in Iraq. Archives of General Psychiatry, 67(6), 614–623. https://doi.org/10.1001/archgenpsychiatry.2010.54

[3] Rytwinski, N. K., Scur, M. D., Feeny, N. C., & Youngstrom, E. A. (2013). The co-occurrence of major depressive disorder among individuals with posttraumatic stress disorder: a meta-analysis: co-occurring PTSD and MDD. Journal of Traumatic Stress, 26(3), 299–309. https://doi.org/10.1002/jts.21814

[4] Flory, J. D., & Yehuda, R. (2015). Comorbidity between post-traumatic stress disorder and major depressive disorder: Alternative explanations and treatment considerations. Dialogues in Clinical Neuroscience, 17(2), 141–150. https://doi.org/10.31887/DCNS.2015.17.2/jflory

[5] Rush, A. J., et al. (2006). Acute and longer-term outcomes in depressed outpatients requiring one or several treatment steps: a STAR*D report. American Journal of Psychiatry, 163(11), 1905–17. https://doi.org/10.1176/appi.ajp.163.11.1905

[6] Watts, B.V., et al. (2013). Meta-analysis of the efficacy of treatments for posttraumatic stress disorder. The Journal of clinical psychiatry, 74(6), e541–50. https://doi.org/10.4088/JCP.12r08225

[7] Holtzheimer, P. E., Russo, J., Zatzick, D., Bundy, C., & Roy-Byrne, P. P. (2005). The impact of comorbid posttraumatic stress disorder on short-term clinical outcome in hospitalized patients with depression. American Journal of Psychiatry, 162(5), 970–976. https://doi.org/10.1176/appi.ajp.162.5.970

[8] Campbell, D. G., et al. (2007). Prevalence of depression–PTSD comorbidity: implications for clinical ractice guidelines and primary care-based interventions. Journal of General Internal Medicine, 22(6), 711–718. https://doi.org/10.1007/s11606-006-0101-4

[9] Green, B.L., et al. (2006). Impact of PTSD comorbidity on one-year outcomes in a depression trial. Journal of Clinical Psychology, 62(7), 815–35. https://doi.org/10.1002/jclp.20279

[10] Hernandez, et al. (2020). Impact of comorbid PTSD on outcome of repetitive transcranial magnetic stimulation (TMS) for veterans with depression. The Journal of Clinical Psychiatry, 81(4), 19m13152. https://doi.org/10.4088/JCP.19m13152

[11] Madore, M. R., et al. (2022). Prefrontal transcranial magnetic stimulation for depression in US military veterans – A naturalistic cohort study in the veterans health administration. Journal of Affective Disorders, 297, 671–678. https://doi.org/10.1016/j.jad.2021.10.025

[12] Carpenter, L. L., et al. (2018). 5 Hz Repetitive transcranial magnetic stimulation for posttraumatic stress disorder comorbid with major depressive disorder. Journal of Affective Disorders, 235, 414–420. https://doi.org/10.1016/j.jad.2018.04.009

[13] Philip, N. S., Doherty, R. A., Faucher, C., Aiken, E., & ‘t Wout-Frank, M. (2022). Transcranial magnetic stimulation for posttraumatic stress disorder and major depression: comparing commonly used clinical protocols. Journal of Traumatic Stress, 35(1), 101–108. https://doi.org/10.1002/jts.22686

[14] Thatcher, R. W. (2011). Neuropsychiatry and quantitative EEG in the 21st Century. Neuropsychiatry, 1(5), 495–514. https://doi.org/10.2217/npy.11.45

[15] Kallioniemi, E., & Daskalakis, Z. J. (2022). Identifying novel biomarkers with TMS-EEG – Methodological possibilities and challenges. Journal of Neuroscience Methods, 377, 109631. https://doi.org/10.1016/j.jneumeth.2022.109631

[16] Arns, M., Drinkenburg, W. H., Fitzgerald, P. B., & Kenemans, J. L. (2012). Neurophysiological predictors of non-response to rTMS in depression. Brain Stimulation, 5(4), 569–576. https://doi.org/10.1016/j.brs.2011.12.003

[17] Valiulis, V., et al. (2012). Electrophysiological differences between high and low frequency rTMS protocols in depression treatment. Acta neurobiologiae experimentalis, 72(3), 283–295.

[18] Noda, Y., et al. (2017). Resting-state EEG gamma power and theta–gamma coupling enhancement following high-frequency left dorsolateral prefrontal rTMS in patients with depression. Clinical Neurophysiology, 128(3), 424–432. https://doi.org/10.1016/j.clinph.2016.12.023

[19] Fuggetta, G., Pavone, E. F., Fiaschi, A., & Manganotti, P. (2008). Acute modulation of cortical oscillatory activities during short trains of high-frequency repetitive transcranial magnetic stimulation of the human motor cortex: A combined EEG and TMS study. Human Brain Mapping, 29(1), 1–13. https://doi.org/10.1002/hbm.20371

[20] Spronk, D., Arns, M., Bootsma, A., van Ruth, R., & Fitzgerald, P. B. (2008). Long term effects of left frontal rTMS on EEG and ERPs in patients with depression. Clinical EEG and Neuroscience, 39(3), 118–124. https://doi.org/10.1177/155005940803900305

[21] Lebiecka, K., et al. (2018). Complexity analysis of EEG data in persons with depression subjected to transcranial magnetic stimulation. Frontiers in Physiology, 9, 1385. https://doi.org/10.3389/fphys.2018.01385

[22] Zandvakili, A., et al. (2019). Use of machine learning in predicting clinical response to transcranial magnetic stimulation in comorbid posttraumatic stress disorder and major depression: A resting state electroencephalography study. Journal of Affective Disorders, 252, 47–54. https://doi.org/10.1016/j.jad.2019.03.077

[23] Zandvakili, A., Swearingen, H. R., & Philip, N. S. (2021). Changes in functional connectivity after theta-burst transcranial magnetic stimulation for post-traumatic stress disorder: A machine-learning study. European Archives of Psychiatry and Clinical Neuroscience, 271(1), 29–37. https://doi.org/10.1007/s00406-020-01172-5

[24] Widge, A. S., et al. (2019). Electroencephalographic biomarkers for treatment response prediction in major depressive illness: A meta-analysis. American Journal of Psychiatry, 176(1), 44–56. https://doi.org/10.1176/appi.ajp.2018.17121358

[25] Jones, S. R. (2016). When brain rhythms aren’t ‘rhythmic’: Implication for their mechanisms and meaning. Current Opinion in Neurobiology, 40, 72–80. https://doi.org/10.1016/j.conb.2016.06.010

[26] van Ede, F., Quinn, A. J., Woolrich, M. W., & Nobre, A. C. (2018). Neural oscillations: sustained rhythms or transient burst-events? Trends in Neurosciences, 41(7), 415–417. https://doi.org/10.1016/j.tins.2018.04.004

[27] Shin, H., Law, R., Tsutsui, S., Moore, C. I., & Jones, S. R. (2017). The rate of transient beta frequency events predicts behavior across tasks and species. ELife, 6, e29086. https://doi.org/10.7554/eLife.29086

[28] Kosciessa, J. Q., Grandy, T. H., Garrett, D. D., & Werkle-Bergner, M. (2020). Single-trial characterization of neural rhythms: Potential and challenges. NeuroImage, 206, 116331. https://doi.org/10.1016/j.neuroimage.2019.116331

[29] Hwang, K., Ghuman, A. S., Manoach, D. S., Jones, S. R., & Luna, B. (2014). Cortical neurodynamics of inhibitory control. Journal of Neuroscience, 34(29), 9551–9561. https://doi.org/10.1523/JNEUROSCI.4889-13.2014

[30] Wagner, J., Wessel, J. R., Ghahremani, A., & Aron, A. R. (2018). Establishing a right frontal beta signature for stopping action in scalp EEG: Implications for testing inhibitory control in other task contexts. Journal of Cognitive Neuroscience, 30(1), 107–118. https://doi.org/10.1162/jocn_a_01183

[31] Seedat, Z. A., et al. (2020). The role of transient spectral ‘bursts’ in functional connectivity: A magnetoencephalography study. NeuroImage, 209, 116537. https://doi.org/10.1016/j.neuroimage.2020.116537

[32] Spitzer, B., & Haegens, S. (2017). Beyond the status quo: A role for beta oscillations in endogenous content (Re)Activation. Eneuro, 4(4), ENEURO.0170-17.2017. https://doi.org/10.1523/ENEURO.0170-17.2017

[33] Lundqvist, M., Herman, P., Warden, M. R., Brincat, S. L., & Miller, E. K. (2018). Gamma and beta bursts during working memory readout suggest roles in its volitional control. Nature Communications, 9(1), 394. https://doi.org/10.1038/s41467-017-02791-8

[34] Bardouille, T., & Bailey, L. (2019). Evidence for age-related changes in sensorimotor neuromagnetic responses during cued button pressing in a large open-access dataset. NeuroImage, 193, 25–34. https://doi.org/10.1016/j.neuroimage.2019.02.065

[35] Hussain, S. J., Cohen, L. G., & Bönstrup, M. (2019). Beta rhythm events predict corticospinal motor output. Scientific Reports, 9(1), 18305. https://doi.org/10.1038/s41598-019-54706-w

[36] Brady, B., Power, L., & Bardouille, T. (2020). Age-related trends in neuromagnetic transient beta burst characteristics during a sensorimotor task and rest in the Cam-CAN open-access dataset. NeuroImage, 222, 117245. https://doi.org/10.1016/j.neuroimage.2020.117245

[37] Levitt, J., et al. (2020). Pain phenotypes classified by machine learning using electroencephalography features. NeuroImage, 223, 117256. https://doi.org/10.1016/j.neuroimage.2020.117256

[38] Sporn, S., Hein, T., & Herrojo Ruiz, M. (2020). Alterations in the amplitude and burst rate of beta oscillations impair reward-dependent motor learning in anxiety. ELife, 9, e50654. https://doi.org/10.7554/eLife.50654

[39] Beam, W., Borckardt, J. J., Reeves, S. T., & George, M. S. (2009). An efficient and accurate new method for locating the F3 position for prefrontal TMS applications. Brain Stimulation, 2(1), 50–54. https://doi.org/10.1016/j.brs.2008.09.006

[40] Rush, A. J., Gullion, C. M., Basco, M. R., Jarrett, R. B., & Trivedi, M. H. (1996). The Inventory of Depressive Symptomatology (IDS): Psychometric properties. Psychological Medicine, 26(3), 477–486. https://doi.org/10.1017/S0033291700035558

[41] Weathers, F.W., Litz, B.T., Keane, T.M., Palmieri, P.A., Marx, B.P., Schnurr, P.P. (2013). The ptsd checklist for dsm-5 (pcl-5). Scale available from the National Center for PTSD at http://www.ptsd.va.gov

[42] Becker, R., et al. (2020). Transient spectral events in resting state MEG predict individual task responses. NeuroImage, 215, 116818. https://doi.org/10.1016/j.neuroimage.2020.116818

[43] Power, L., & Bardouille, T. (2021). Age-related trends in the cortical sources of transient beta bursts during a sensorimotor task and rest. NeuroImage, 245, 118670. https://doi.org/10.1016/j.neuroimage.2021.118670

[44] Gómez, C. M., Angulo-Ruíz, B. Y., Muñoz, V., & Rodriguez-Martínez, E. I. (2022). Activation-Inhibition dynamics of the oscillatory bursts of the human EEG during resting state. The macroscopic temporal range of few seconds. Cognitive Neurodynamics, 16(3), 591–608. https://doi.org/10.1007/s11571-021-09742-6

[45] Julkunen, P., Kimiskidis, V. K., & Belardinelli, P. (2022). Bridging the gap: TMS-EEG from lab to clinic. Journal of Neuroscience Methods, 369, 109482. https://doi.org/10.1016/j.jneumeth.2022.109482

[46] Price, G. W., Lee, J. W., Garvey, C., & Gibson, N. (2008). Appraisal of sessional EEG features as a correlate of clinical changes in an rTMS treatment of depression. Clinical EEG and Neuroscience, 39(3), 131–138. https://doi.org/10.1177/155005940803900307

[47] Widge, A. S., Avery, D. H., & Zarkowski, P. (2013). Baseline and treatment-emergent EEG biomarkers of antidepressant medication response do not predict response to repetitive transcranial magnetic stimulation. Brain Stimulation, 6(6), 929–931. https://doi.org/10.1016/j.brs.2013.05.001

[48] Petrosino, N. J., Zandvakili, A., Carpenter, L. L., & Philip, N. S. (2018). Pilot testing of peak alpha frequency stability during repetitive transcranial magnetic stimulation. Frontiers in Psychiatry, 9, 605. https://doi.org/10.3389/fpsyt.2018.00605

[49] Kito, S., et al. (2017). Transcranial magnetic stimulation modulates resting EEG functional connectivity between the left dorsolateral prefrontal cortex and limbic regions in medicated patients with treatment-resistant depression. The Journal of Neuropsychiatry and Clinical Neurosciences, 29(2), 155–159. https://doi.org/10.1176/appi.neuropsych.15120419

[50] Bailey, N. W., et al. (2018). Responders to rTMS for depression show increased fronto-midline theta and theta connectivity compared to non-responders. Brain Stimulation, 11(1), 190–203. https://doi.org/10.1016/j.brs.2017.10.015

[51] Olejarczyk, E., et al. (2020). The impact of repetitive transcranial magnetic stimulation on functional connectivity in major depressive disorder and bipolar disorder evaluated by directed transfer function and indices based on graph theory. International Journal of Neural Systems, 30(04), 2050015. https://doi.org/10.1142/S012906572050015X

[52] Silverstein, W. K., et al. (2015). Neurobiological predictors of response to dorsolateral prefrontal cortex repetitive transcranial magnetic stimulation in depression: a Systematic Review. Depression and anxiety, 32(12), 871–891. https://doi.org/10.1002/da.22424

[53] Hoogendam, J. M., Ramakers, G. M. J., & Di Lazzaro, V. (2010). Physiology of repetitive transcranial magnetic stimulation of the human brain. Brain Stimulation, 3(2), 95–118. https://doi.org/10.1016/j.brs.2009.10.005

[54] Philip, N. S., et al. (2018). Network mechanisms of clinical response to transcranial magnetic stimulation in posttraumatic stress disorder and major depressive disorder. Biological Psychiatry, 83(3), 263–272. https://doi.org/10.1016/j.biopsych.2017.07.021

[55] Levy, B. J., & Wagner, A. D. (2011). Cognitive control and right ventrolateral prefrontal cortex: reflexive reorienting, motor inhibition, and action updating. Annals of the New York Academy of Sciences, 1224(1), 40–62. https://doi.org/10.1111/j.1749-6632.2011.05958

[56] Whitton, A. E., et al. (2018). Electroencephalography source functional connectivity reveals abnormal high-frequency communication among large-scale functional networks in depression. Biological Psychiatry: Cognitive Neuroscience and Neuroimaging, 3(1), 50–58. https://doi.org/10.1016/j.bpsc.2017.07.001

[57] Roh, S.-C., Park, E.-J., Shim, M., & Lee, S.-H. (2016). EEG beta and low gamma power correlates with inattention in patients with major depressive disorder. Journal of Affective Disorders, 204, 124–130. https://doi.org/10.1016/j.jad.2016.06.033

[58] Popescu, M., Popescu, E.-A., DeGraba, T. J., & Hughes, J. D. (2020). Altered modulation of beta band oscillations during memory encoding is predictive of lower subsequent recognition performance in post-traumatic stress disorder. NeuroImage: Clinical, 25, 102154. https://doi.org/10.1016/j.nicl.2019.102154

[59] Cohen, J. E., et al. (2013). Emotional brain rhythms and their impairment in post-traumatic patients. Human Brain Mapping, 34(6), 1344–1356. https://doi.org/10.1002/hbm.21516

[60] Sacchet, M. D., et al. (2015). Attention drives synchronization of alpha and beta rhythms between right inferior frontal and primary sensory neocortex. Journal of Neuroscience, 35(5), 2074–2082. https://doi.org/10.1523/JNEUROSCI.1292-14.2015

[61] Swann, N., et al. (2009). Intracranial EEG reveals a time- and frequency-specific role for the right inferior frontal gyrus and primary motor cortex in stopping initiated responses. The Journal of neuroscience: the official journal of the Society for Neuroscience, 29(40), 12675–12685. https://doi.org/10.1523/JNEUROSCI.3359-09.2009

[62] Sundby, K. K., Jana, S., & Aron, A. R. (2021). Double-blind disruption of right inferior frontal cortex with TMS reduces right frontal beta power for action stopping. Journal of Neurophysiology, 125(1), 140–153. https://doi.org/10.1152/jn.00459.2020

[63] Wessel, J. R. (2020). Β-Bursts reveal the trial-to-trial dynamics of movement initiation and cancellation. The Journal of Neuroscience, 40(2), 411–423. https://doi.org/10.1523/JNEUROSCI.1887-19.2019

[64] Jana, S., Hannah, R., Muralidharan, V., & Aron, A. R. (2020). Temporal cascade of frontal, motor and muscle processes underlying human action-stopping. ELife, 9, e50371. https://doi.org/10.7554/eLife.50371

[65] Hannah, R., Muralidharan, V., Sundby, K. K., & Aron, A. R. (2020). Temporally-precise disruption of prefrontal cortex informed by the timing of beta bursts impairs human action-stopping. NeuroImage, 222, 117222. https://doi.org/10.1016/j.neuroimage.2020.117222

[66] Errington, S. P., Woodman, G. F., & Schall, J. D. (2020). Dissociation of medial frontal β-bursts and executive control. The Journal of Neuroscience, 40(48), 9272–9282. https://doi.org/10.1523/JNEUROSCI.2072-20.2020

[67] Enz, N., Ruddy, K. L., Rueda-Delgado, L. M., & Whelan, R. (2021). Volume of β-Bursts, But Not Their Rate, Predicts Successful Response Inhibition. The Journal of Neuroscience, 41(23), 5069–5079. https://doi.org/10.1523/JNEUROSCI.2231-20.2021

[68] Sherman, M. A., et al. (2016). Neural mechanisms of transient neocortical beta rhythms: Converging evidence from humans, computational modeling, monkeys, and mice. Proceedings of the National Academy of Sciences, 113(33). https://doi.org/10.1073/pnas.1604135113

[69] Neymotin, S. A., et al. (2020). Human neocortical neurosolver (HNN), a new software tool for interpreting the cellular and network origin of human MEG/EEG data. ELife, 9, e51214. https://doi.org/10.7554/eLife.51214

[70] Law, R. G., et al. (2022). Thalamocortical mechanisms regulating the relationship between transient beta events and human tactile perception. Cerebral Cortex, 32(4), 668–688. https://doi.org/10.1093/cercor/bhab221

[71] Bonaiuto, J. J., et al. (2021). Laminar dynamics of high amplitude beta bursts in human motor cortex. NeuroImage, 242, 118479. https://doi.org/10.1016/j.neuroimage.2021.118479

[72] Scheyltjens, I., & Arckens, L. (2016). The current status of somatostatin-interneurons in inhibitory control of brain function and plasticity. Neural Plasticity, 2016, 1–20. https://doi.org/10.1155/2016/8723623

[73] Williams, L. E., & Holtmaat, A. (2019). Higher-order thalamocortical inputs gate synaptic long-term potentiation via disinhibition. Neuron, 101(1), 91- 102.e4. https://doi.org/10.1016/j.neuron.2018.10.049

[74] Benjamini Y, Hochberg Y. Controlling the False Discovery Rate: A Practical and Powerful Approach to Multiple Testing. Journal of the Royal Statistical Society: Series B (Methodological) 1995;57:289–300. https://doi.org/10.1111/j.2517-6161.1995.tb02031.x

