## Supplementary material for "Fronto-central resting-state 15-29Hz transient beta events change with therapeutic transcranial magnetic stimulation for posttraumatic stress disorder and major depressive disorder": Manuscript

Supplementary Information

### Threshold Sensitivity Analysis Supports a 6X FOM Event Detection Threshold

We performed several analyses as in Shin et al. (2017) to provide objective measurements supporting the choice of a 6X factor of the median (FOM) power threshold for detecting events (Fig. S1). For a range of 0.25-16 FOM threshold values, and for each frequency band, we computed the percent area of the suprathreshold region above this cutoff in each epoch's spectrogram (Fig. S1.a inset) and calculated its correlation with the epoch's mean power averaged across frequency and time (Fig. S1.a). The correlation between mean power and suprathreshold area was highest around the 6X FOM power cutoff across frequency bands, shown as a dotted line in Fig. S1.a, consistent with Shin et al 2017. Furthermore, the inverse cumulative density function (1-CDF) of local maxima across all epochs shows the percent captured by each potential FOM cutoff (Fig. S1.b). The dotted line shows that about 20% of all local maxima are registered as events using a 6X FOM cutoff. Based on these analyses, we used a 6X FOM cutoff to detect events for each frequency band.

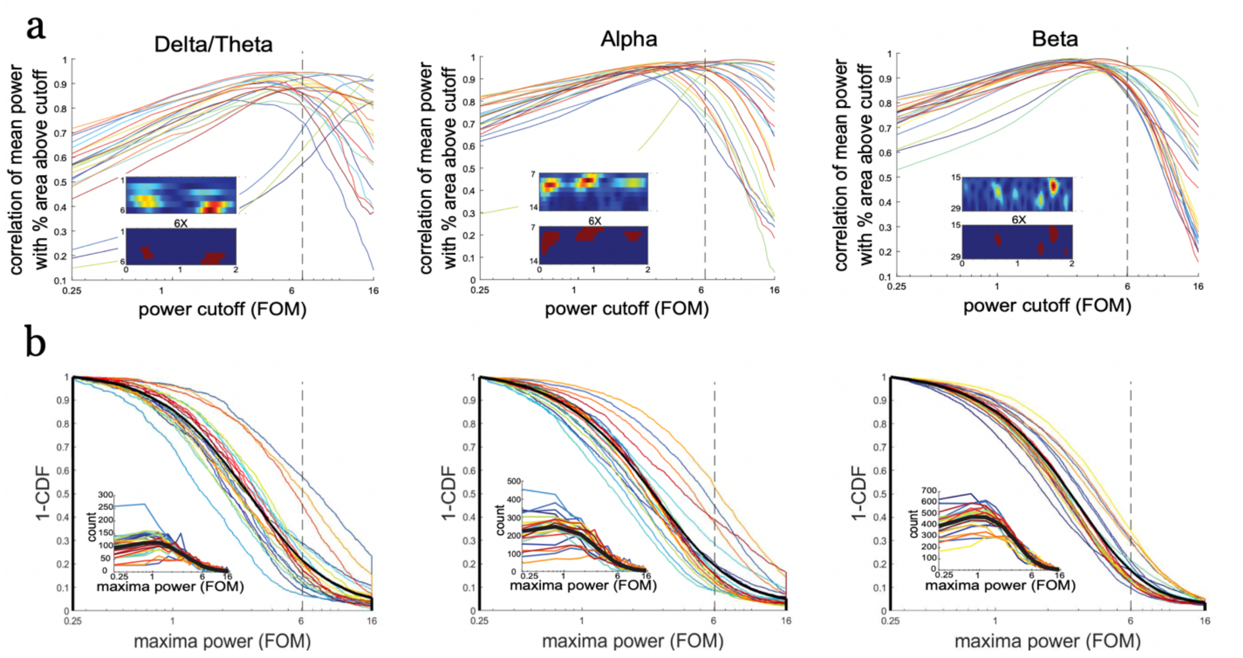

**Figure S1** Events defined by a threshold set at 6X (factor of the median, FOM) power consistently show highest correlation with time-averaged power across the entire time-frequency epoch (example data from Fpz electrode). (a) Pearson's correlation coefficient between average power across entire time-frequency and the percent area (percentage of non-zero pixels in the spectrogram) above FOM thresholds plotted on a log scale, for each BOI (colored lines for individual patients; black line for average across patients). Inset in A shows an example spectrogram (top) and the percent area above a 6X FOM cutoff (below). (b) Distribution of all local maxima peak power values in non-averaged spectrograms. 1-CDF shows the proportion of local maxima above each threshold (colored lines for individual patients; black line for aggregate of all local maxima across patients). Inset histograms show the same data.

### Additional Event Feature Information

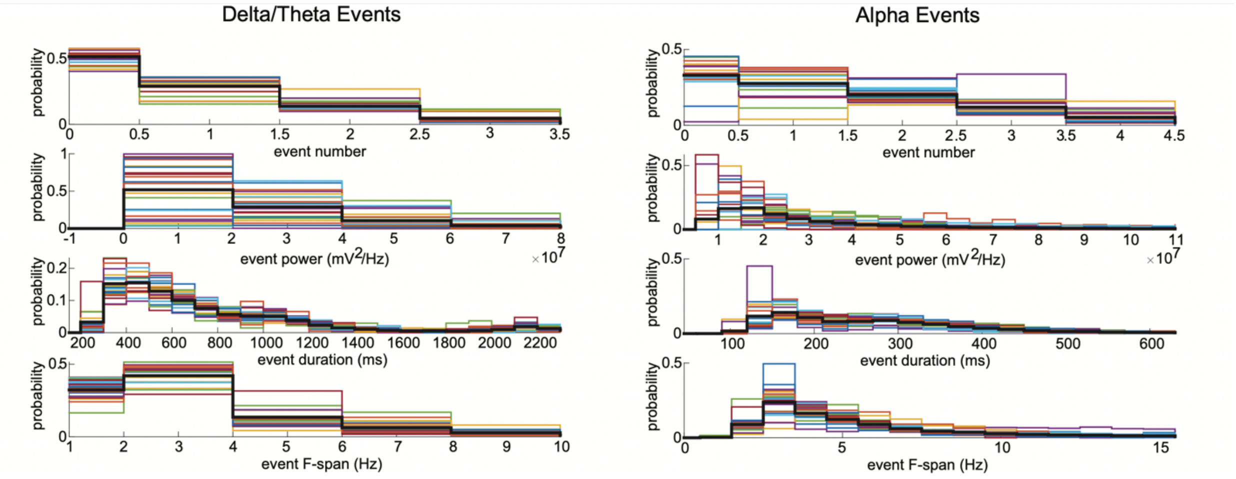

**Figure S2** Histograms showing the distribution of all delta/theta (left) and alpha (right) event characteristic measurements for individual patients (colored lines) and the average (black line) from the Fpz electrode.

| | **Delta/Theta** | | | | **Alpha** | | | | **Beta** | | | |
|---|---|---|---|---|---|---|---|---|---|---|---|---|
| | Number | Power | Duration | F-span | Number | Power | Duration | F-span | Number | Power | Duration | F-span |
| **Fp1** | 0.74±0.04 | 3.02±0.57 E+07 | 856.31 ±21.07 | 3.20±0.13 | 1.40±0.06 | 3.88±0.60 E+07 | 290.22 ±9.26 | 5.88±0.33 | 2.52±0.09 | 2.56±0.31 E+07 | 138.95 ±2.26 | 10.03±0.21 |
| **Fpz** | 0.75±0.04 | 3.00±0.51 E+07 | 872.03 ±20.00 | 3.13±0.12 | 1.36±0.06 | 3.93±0.58 E+07 | 295.92 ±10.52 | 5.67±0.33 | 2.47±0.08 | 2.20±0.16 E+07 | 137.95 ±1.40 | 10.07±0.27 |
| **Fp2** | 0.75±0.04 | 2.95±0.51 E+07 | 886.06 ±22.81 | 3.12±0.16 | 1.40±0.06 | 3.63±0.50 E+07 | 297.00 ±7.75 | 5.76±0.32 | 2.51±0.08 | 2.24±0.17 E+07 | 138.14 ±1.44 | 9.97±0.25 |
| **Fz** | 0.65±0.03 | 2.63±0.41 E+07 | 829.62 ±34.44 | 3.06±0.12 | 1.28±0.05 | 3.71±0.46 E+07 | 296.72 ±7.40 | 5.39±0.18 | 2.34±0.06 | 2.19±0.12 E+07 | 137.62 ±1.87 | 9.68±0.12 |
| **Cz** | 0.67±0.03 | 1.78±0.26 E+07 | 903.96 ±29.91 | 2.92±0.13 | 1.17±0.03 | 2.67±0.42 E+07 | 302.66 ±6.98 | 5.03±0.10 | 2.17±0.05 | 2.08±0.06 E+07 | 136.30 ±1.07 | 9.47±0.07 |
| **Pz** | 0.63±0.03 | 1.30±0.13 E+07 | 940.71 ±22.46 | 2.98±0.31 | 1.13±0.04 | 2.12±0.29 E+07 | 295.25 ±4.69 | 5.09±0.09 | 2.15±0.05 | 2.08±0.09 E+07 | 135.73 ±1.87 | 9.48±0.07 |
| **Oz** | 0.68±0.04 | 2.02±0.32 | 901.99 | 3.16±0.20 | 1.26±0.05 | 3.43±0.61 | 299.14 | 5.44±0.28 | 2.28±0.06 | 2.22±0.22 | 134.40 | 9.95±0.23 |

| | | | | | | | | | | | | |
|---|---|---|---|---|---|---|---|---|---|---|---|---|
| | | E+07 | ±28.78 | | | E+07 | ±7.59 | | | E+07 | ±1.16 | |
| **F3** | 0.65±0.02 | 2.63±0.35 E+07 | 886.17 ±22.55 | 3.12±0.13 | 1.27±0.06 | 3.64±0.44 E+07 | 285.96 ±7.59 | 5.67±0.31 | 2.38±0.08 | 2.28±0.15 E+07 | 135.84 ±1.29 | 9.94±0.20 |
| **AVERAGE** | 0.69±0.03 | 2.41±0.38 E+07 | 884.60 ±25.25 | 3.09±0.16 | 1.28±0.05 | 3.38±0.49 E+07 | 295.36 ±7.72 | 5.49±0.24 | 2.35±0.07 | 2.23±0.16 E+07 | 136.87 ±1.54 | 9.82±0.18 |

**Table S1** Average event features across patients (mean ± standard error), for each electrode and BOI. Number (counts/2-sec epoch), power ($mv^2$/Hz), duration (ms), and frequency Span (Hz). Average values across electrodes are also reported.

### Further Illustration of the Changes in Beta Event Features

To further illustrate the relationship between beta event features and symptom improvement, we sorted patients according to the magnitude of their symptom reduction following rTMS and grouped the top third (those with greatest post-treatment improvement) and bottom third (those with least improvement or worsening symptoms) (n=7 per group) and plotted beta event features from these extremes.

For each group, we first constructed boxplots of the event feature changes from pre- to post-treatment (Fig. S3). On average, frontal beta event duration increased within the top third patient group showing improved IDS-SR scores, and decreased within the bottom third group, while frequency spans showed the opposite effect. (Fig. S3, left and middle). Central beta event maxima power decreased on average in the top third group showing improved PCL-5 scores and increased in the bottom third group (Fig. S3, right). The pre- to post- treatment percent change of each feature was significantly different when comparing across groups (frequency span p=0.003, duration p=0.044, maximum power p=0.025), but did not reach within group significance due to low sample sizes in this illustration.

We next illustrated each event's time-frequency representation using the boundaries of the supra-threshold time-frequency region containing the event in each group (Fig. S4). To visualize changes in frontal electrode beta event duration and frequency span and central electrode beta event maxima power that occur with symptom improvement, we aligned events at the time and frequency of power maximum, and then averaged the normalized time-frequency response of the region for each event above half of its peak power for each patient. We averaged the spectrograms and created separate spectrograms for the pre- and post-treatment conditions for each group and electrode. Consistent with the computation of duration and frequency span for edge cases, if the event was at the edge in time (2-second epoch edges) and/or frequency (frequency band edges), the region was extended by doubling the side that was not cut by the edge.  All spectrograms are averaged over an equal number of events during pre- and post-treatment sessions.

In the Fpz spectrograms (Fig. S4, left), the duration can be visualized increasing for the top third and decreasing for the bottom third group, and the frequency span can be visualized decreasing in the upper third and increasing in the bottom third after treatment. In the Cz spectrograms (Fig. S4, right), the FOM maxima power can be visualized decreasing in the upper third and increasing in the bottom third after treatment.

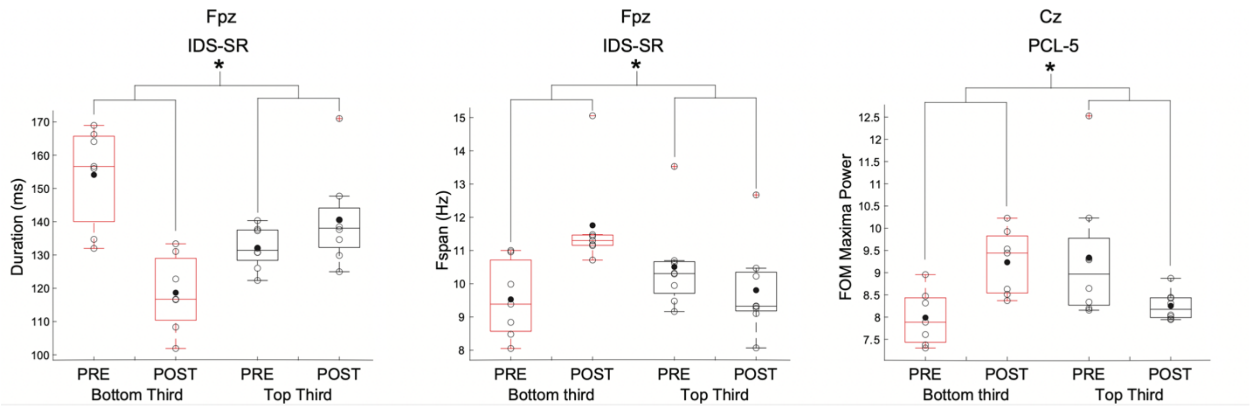

**Figure S3** Boxplots showing the pre- to post-treatment averages for beta event duration (left), frequency span (middle), and FOM maxima power (right) for patients grouped by clinical response into the top and bottom third of IDS-SR or PCL-5 scored symptom improvements. Boxplots mark the range, 25th percentile, 75th percentile, and median, including individual patients (o)

and the mean (•). (*) Denotes significant corrected p values for t-tests comparing percent change pre- to post-treatment between groups.

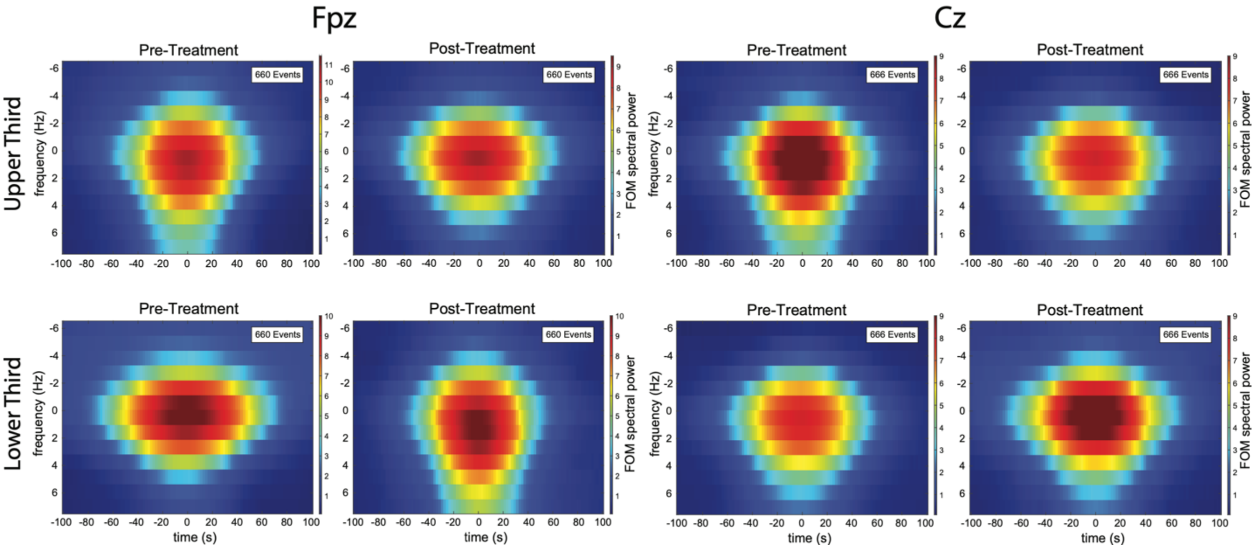

**Figure S4** Beta event spectrograms depicting averaged normalized spectral power in a 200ms time window aligned at the maxima time point of each Fpz and Cz beta event pre- and post-treatment for the upper third and lower third of all patients grouped by clinical scores from the IDS-SR and PCL-5. The heat map (color bar) shows FOM power values. The number of events included (legend) constitutes a random sample of an equal number of events across patients and sessions.

| | | Delta/Theta | | | | Alpha | | | | Beta | | | |
|---|---|---|---|---|---|---|---|---|---|---|---|---|---|
| | | Number | Power | Duration | F-span | Number | Power | Duration | F-span | Number | Power | Duration | F-span |
| F3 | pc | 14.4272 | 8.4230 | -6.9603 | 1.1140 | 2.8883 | -2.8523 | 5.6619 | -3.0625 | -15.2030 | -2.1506 | 1.8749 | -0.7975 |
| | p-value (u) | 0.5727 | 0.5325 | 0.3510 | 0.8836 | 0.8888 | 0.7992 | 0.2544 | 0.4720 | 0.1246 | 0.6867 | 0.5465 | 0.5455 |
| | p-value (c) | 0.9261 | 0.9261 | 0.9261 | 0.9509 | 0.9509 | 0.9509 | 0.9261 | 0.9261 | 0.7974 | 0.9421 | 0.9261 | 0.9261 |
| Fp1 | pc | -42.0853 | 13.7425 | 4.2108 | -16.3493 | -35.8694 | -15.0661 | -8.1745 | 0.7220 | -31.4788 | -7.5675 | -6.4375 | 1.6072 |
| | p-value (u) | 0.0422 | 0.6097 | 0.6367 | 0.0111 | 0.0566 | 0.1740 | 0.1689 | 0.8923 | 0.0398 | 0.1145 | 0.1838 | 0.5985 |
| | p-value (c) | 0.7974 | 0.9261 | 0.9261 | 0.6656 | 0.7974 | 0.8319 | 0.8319 | 0.9509 | 0.7974 | 0.7974 | 0.8319 | 0.9261 |
| Fpz | pc | -48.9224 | 14.0650 | 4.7231 | -14.2205 | -42.6965 | -17.0254 | -9.4530 | 1.0509 | -12.7458 | -2.3667 | -6.6593 | 3.0398 |
| | p-value (u) | 0.0208 | 0.5677 | 0.5276 | 0.0635 | 0.0143 | 0.0994 | 0.1215 | 0.8712 | 0.3038 | 0.5698 | 0.1216 | 0.4052 |
| | p-value (c) | 0.6656 | 0.9261 | 0.9261 | 0.7974 | 0.6656 | 0.7974 | 0.7974 | 0.9509 | 0.9261 | 0.9261 | 0.7974 | 0.9261 |
| Fp2 | pc | -18.0897 | 23.7653 | 12.5119 | -14.1929 | -25.0418 | -7.3031 | -10.2078 | 3.2946 | -11.5265 | -4.0858 | -2.7384 | 2.0515 |
| | p-value (u) | 0.4235 | 0.4542 | 0.1967 | 0.0877 | 0.1955 | 0.5089 | 0.0885 | 0.6183 | 0.4679 | 0.3337 | 0.4776 | 0.5501 |
| | p-value (c) | 0.9261 | 0.9261 | 0.8319 | 0.7974 | 0.8319 | 0.9261 | 0.7974 | 0.9261 | 0.9261 | 0.9261 | 0.9261 | 0.9261 |
| Fz | pc | -13.8428 | 15.0516 | 3.5839 | 7.1220 | 4.1034 | -7.4342 | -3.4788 | 1.4363 | -3.4511 | -1.8216 | 0.6568 | 0.7486 |
| | p-value (u) | 0.4787 | 0.2897 | 0.7680 | 0.3930 | 0.8685 | 0.4972 | 0.4915 | 0.6966 | 0.7985 | 0.7467 | 0.8499 | 0.6921 |
| | p-value (c) | 0.9261 | 0.9261 | 0.9509 | 0.9261 | 0.9509 | 0.9261 | 0.9261 | 0.9421 | 0.9509 | 0.9509 | 0.9509 | 0.9421 |
| Cz | pc | 12.1008 | 16.2337 | -4.7248 | 0.7584 | -6.6255 | 15.2393 | -1.5168 | -0.6759 | 0.6230 | -3.7774 | 1.7552 | 0.2126 |
| | p-value (u) | 0.6986 | 0.3280 | 0.5883 | 0.9431 | 0.7983 | 0.5225 | 0.8028 | 0.8781 | 0.9473 | 0.3176 | 0.5771 | 0.8861 |
| | p-value (c) | 0.9421 | 0.9261 | 0.9261 | 0.9779 | 0.9509 | 0.9261 | 0.9509 | 0.9509 | 0.9779 | 0.9261 | 0.9261 | 0.9509 |
| Pz | pc | -28.7785 | -0.1271 | -12.2075 | -8.7312 | -7.9749 | -0.3215 | -3.4774 | 2.2226 | -2.1175 | -4.6499 | -0.0536 | 0.6227 |
| | p-value (u) | 0.1964 | 0.9858 | 0.1141 | 0.5848 | 0.7351 | 0.5197 | 0.5578 | 0.6102 | 0.8461 | 0.3672 | 0.9854 | 0.6300 |
| | p-value (c) | 0.8319 | 0.9858 | 0.7974 | 0.9261 | 0.9509 | 0.9261 | 0.9261 | 0.9261 | 0.9509 | 0.9261 | 0.9858 | 0.9261 |

| | | | | | | | | | | | | | |
|---|---|---|---|---|---|---|---|---|---|---|---|---|---|
| Oz | pc | 21.2037 | 18.1047 | -2.9666 | -0.2505 | 29.5296 | 22.3234 | 2.4136 | 0.7528 | 15.2927 | 4.8311 | 0.5455 | 1.0226 |
| | p-value (u) | 0.5514 | 0.1733 | 0.6677 | 0.9790 | 0.3457 | 0.2122 | 0.7164 | 0.9014 | 0.1993 | 0.6015 | 0.8822 | 0.7115 |
| | p-value (c) | 0.9261 | 0.8319 | 0.9421 | 0.9858 | 0.9261 | 0.8488 | 0.9421 | 0.9509 | 0.8319 | 0.9261 | 0.9509 | 0.9421 |

**Table S2** Event feature comparisons from pre- to post-treatment. T-tests compared pre- and post-treatment feature values, computed separately for each frequency band and each electrode. Corrected (c) values using the Benjamini–Hochberg procedure and uncorrected (u) p values reported, none trending (corrected value $0.05 < p < 0.08$); or significant (corrected $p < 0.05$). Percent change (pc) values are calculated as 100*(post-pre)/pre.

**a**

| **IDS-SR** | | **Delta/Theta** | | | | **Alpha** | | | | **Beta** | | | |
|---|---|---|---|---|---|---|---|---|---|---|---|---|---|
| | | Number | Power | Duration | F-span | Number | Power | Duration | F-span | Number | Power | Duration | F-span |
| **F3** | r | 0.2722 | 0.3587 | -0.3921 | 0.1845 | 0.3099 | 0.1834 | 0.0602 | 0.0330 | -0.1867 | 0.4716 | 0.0555 | 0.0846 |
| | **$r^2$** | 0.0741 | 0.1286 | 0.1537 | 0.0340 | 0.0960 | 0.0336 | 0.0036 | 0.0011 | 0.0349 | 0.2224 | 0.0031 | 0.0072 |
| | p-value (u) | 0.2090 | 0.0929 | 0.0643 | 0.3995 | 0.1501 | 0.4021 | 0.7849 | 0.8810 | 0.3937 | 0.0231 | 0.8013 | 0.7010 |
| | p-value (c) | 0.5172 | 0.4694 | 0.4115 | 0.6688 | 0.5172 | 0.6688 | 0.8831 | 0.9193 | 0.6688 | 0.1721 | 0.8842 | 0.8489 |

| | | | | | | | | | | | | | |
|---|---|---|---|---|---|---|---|---|---|---|---|---|---|
| **Fp1** | r | -0.0585 | 0.1490 | 0.2233 | -0.0827 | -0.1472 | -0.2076 | -0.1128 | -0.1581 | -0.1211 | -0.2725 | ***-0.6620*** | 0.5502 |
| | **$r^2$** | 0.0034 | 0.0222 | 0.0499 | 0.0068 | 0.0217 | 0.0431 | 0.0127 | 0.0250 | 0.0147 | 0.0743 | ***0.4382*** | 0.3027 |
| | p-value (u) | 0.7911 | 0.4973 | 0.3057 | 0.7074 | 0.5026 | 0.3419 | 0.6082 | 0.4713 | 0.5821 | 0.2084 | ***0.0006*** | 0.0065 |
| | p-value (c) | 0.8831 | 0.7244 | 0.5989 | 0.8489 | 0.7244 | 0.6193 | 0.7998 | 0.7182 | 0.7977 | 0.5172 | ***0.0144*** | 0.0780 |
| **Fpz** | r | -0.1072 | 0.2375 | 0.4829 | -0.3286 | -0.0715 | -0.1774 | -0.2521 | 0.1294 | 0.2822 | -0.2079 | ***-0.6616*** | ***0.6854*** |
| | **$r^2$** | 0.0115 | 0.0564 | 0.2332 | 0.1080 | 0.0051 | 0.0315 | 0.0636 | 0.0167 | 0.0796 | 0.0432 | ***0.4378*** | ***0.4698*** |
| | p-value (u) | 0.6262 | 0.2751 | 0.0196 | 0.1258 | 0.7459 | 0.4180 | 0.2458 | 0.5563 | 0.1920 | 0.3411 | ***0.0006*** | ***0.0003*** |
| | p-value (c) | 0.8124 | 0.5709 | 0.1721 | 0.4866 | 0.8735 | 0.6688 | 0.5483 | 0.7854 | 0.5172 | 0.6193 | ***0.0144*** | ***0.0144*** |
| **Fp2** | r | -0.1247 | 0.2271 | 0.2080 | -0.0366 | -0.0596 | -0.1462 | -0.3685 | 0.3238 | 0.2565 | -0.1151 | ***-0.6296*** | ***0.6625*** |
| | **$r^2$** | 0.0155 | 0.0516 | 0.0433 | 0.0013 | 0.0035 | 0.0214 | 0.1358 | 0.1048 | 0.0658 | 0.0132 | ***0.3964*** | ***0.4389*** |
| | p-value (u) | 0.5709 | 0.2973 | 0.3409 | 0.8684 | 0.7872 | 0.5056 | 0.0836 | 0.1318 | 0.2375 | 0.6011 | ***0.0013*** | ***0.0006*** |
| | p-value (c) | 0.7943 | 0.5946 | 0.6193 | 0.9161 | 0.8831 | 0.7244 | 0.4459 | 0.4866 | 0.5483 | 0.7998 | ***0.0250*** | ***0.0144*** |
| **Fz** | r | 0.1043 | 0.2766 | 0.0492 | 0.3029 | 0.2397 | -0.0009 | -0.1876 | 0.1811 | 0.2685 | **0.5710** | 0.1186 | 0.0832 |
| | **$r^2$** | 0.0109 | 0.0765 | 0.0024 | 0.0918 | 0.0574 | 0.0000 | 0.0352 | 0.0328 | 0.0721 | **0.3260** | 0.0141 | 0.0069 |
| | p-value (u) | 0.6356 | 0.2014 | 0.8235 | 0.1600 | 0.2707 | 0.9967 | 0.3914 | 0.4083 | 0.2155 | **0.0044** | 0.5900 | 0.7059 |
| | p-value (c) | 0.8136 | 0.5172 | 0.8969 | 0.5172 | 0.5709 | 0.9967 | 0.6688 | 0.6688 | 0.5172 | **0.0603** | 0.7977 | 0.8489 |
| **Cz** | r | 0.2788 | 0.4239 | -0.2999 | 0.2819 | 0.1691 | 0.3775 | 0.2783 | 0.0174 | 0.0369 | ***0.5902*** | 0.2881 | 0.0714 |
| | **$r^2$** | 0.0777 | 0.1797 | 0.0900 | 0.0795 | 0.0286 | 0.1425 | 0.0775 | 0.0003 | 0.0014 | ***0.3484*** | 0.0830 | 0.0051 |
| | p-value (u) | 0.1976 | 0.0438 | 0.1644 | 0.1925 | 0.4405 | 0.0758 | 0.1985 | 0.9373 | 0.8672 | ***0.0030*** | 0.1825 | 0.7461 |
| | p-value (c) | 0.5172 | 0.3003 | 0.5172 | 0.5172 | 0.6932 | 0.4365 | 0.5172 | 0.9572 | 0.9161 | ***0.0480*** | 0.5172 | 0.8735 |
| **Pz** | r | -0.2690 | 0.4744 | -0.3237 | -0.1551 | 0.0481 | -0.0267 | -0.1650 | 0.2578 | 0.2493 | 0.5091 | 0.0135 | 0.2355 |
| | **$r^2$** | 0.0723 | 0.2250 | 0.1048 | 0.0240 | 0.0023 | 0.0007 | 0.0272 | 0.0665 | 0.0622 | 0.2591 | 0.0002 | 0.0554 |
| | p-value (u) | 0.2146 | 0.0222 | 0.1318 | 0.4798 | 0.8315 | 0.9060 | 0.4631 | 0.2467 | 0.2513 | 0.0131 | 0.9514 | 0.2795 |

| | | | | | | | | | | | | | |
|---|---|---|---|---|---|---|---|---|---|---|---|---|---|
| | p-value (c) | 0.5172 | 0.1721 | 0.4866 | 0.7197 | 0.8969 | 0.9352 | 0.7171 | 0.5483 | 0.5483 | 0.1397 | 0.9614 | 0.5709 |
| **Oz** | r | 0.3369 | 0.4711 | -0.0656 | 0.3461 | 0.3497 | 0.3021 | 0.1786 | -0.2157 | 0.3756 | 0.3311 | 0.0964 | -0.0904 |
| | **$r^2$** | 0.1135 | 0.2220 | 0.0043 | 0.1198 | 0.1223 | 0.0913 | 0.0319 | 0.0465 | 0.1411 | 0.1097 | 0.0093 | 0.0082 |
| | p-value (u) | 0.1160 | 0.0233 | 0.7660 | 0.1058 | 0.1019 | 0.1612 | 0.4149 | 0.3230 | 0.0773 | 0.1227 | 0.6618 | 0.6817 |
| | p-value (c) | 0.4866 | 0.1721 | 0.8831 | 0.4837 | 0.4837 | 0.5172 | 0.6688 | 0.6193 | 0.4365 | 0.4866 | 0.8360 | 0.8489 |

# b

| **PCL-5** | | **Delta/Theta** | | | | **Alpha** | | | | **Beta** | | | |
|---|---|---|---|---|---|---|---|---|---|---|---|---|---|
| | | Number | Power | Duration | F-span | Number | Power | Duration | F-span | Number | Power | Duration | F-span |
| **F3** | r | 0.0090 | 0.0986 | -0.4812 | -0.0398 | 0.0491 | -0.0223 | -0.0722 | 0.0135 | -0.1810 | 0.3653 | -0.1135 | 0.2051 |
| | **$r^2$** | 0.0001 | 0.0097 | 0.2316 | 0.0016 | 0.0024 | 0.0005 | 0.0052 | 0.0002 | 0.0328 | 0.1334 | 0.0129 | 0.0421 |
| | p-value (u) | 0.9674 | 0.6546 | 0.0201 | 0.8571 | 0.8240 | 0.9197 | 0.7433 | 0.9511 | 0.4085 | 0.0865 | 0.6062 | 0.3478 |
| | p-value (c) | 0.9674 | 0.8925 | 0.2144 | 0.9549 | 0.9525 | 0.9674 | 0.9052 | 0.9674 | 0.7577 | 0.4848 | 0.8817 | 0.7267 |
| **Fp1** | r | -0.0885 | 0.1902 | 0.1357 | -0.2246 | -0.2138 | -0.2130 | -0.1216 | -0.2228 | -0.2018 | -0.3192 | -0.5555 | 0.3937 |
| | **$r^2$** | 0.0078 | 0.0362 | 0.0184 | 0.0504 | 0.0457 | 0.0454 | 0.0148 | 0.0496 | 0.0407 | 0.1019 | 0.3086 | 0.1550 |
| | p-value (u) | 0.6880 | 0.3847 | 0.5370 | 0.3029 | 0.3274 | 0.3291 | 0.5804 | 0.3070 | 0.3558 | 0.1376 | 0.0059 | 0.0631 |
| | p-value (c) | 0.8925 | 0.7577 | 0.8183 | 0.7188 | 0.7267 | 0.7267 | 0.8624 | 0.7188 | 0.7267 | 0.5624 | 0.1416 | 0.4848 |
| **Fpz** | r | -0.0988 | 0.2636 | 0.3754 | -0.3223 | -0.0769 | -0.1597 | -0.1851 | 0.0720 | 0.1508 | -0.1829 | -0.5621 | 0.5900 |
| | **$r^2$** | 0.0098 | 0.0695 | 0.1409 | 0.1039 | 0.0059 | 0.0255 | 0.0343 | 0.0052 | 0.0227 | 0.0335 | 0.3160 | 0.3481 |
| | p-value (u) | 0.6538 | 0.2243 | 0.0776 | 0.1336 | 0.7272 | 0.4666 | 0.3977 | 0.7441 | 0.4923 | 0.4035 | 0.0052 | 0.0030 |
| | p-value (c) | 0.8925 | 0.6813 | 0.4848 | 0.5624 | 0.9052 | 0.7953 | 0.7577 | 0.9052 | 0.7991 | 0.7577 | 0.1416 | 0.1416 |
| **Fp2** | r | -0.1548 | 0.2631 | -0.0381 | -0.0105 | -0.0522 | -0.0921 | -0.2432 | 0.2065 | 0.2747 | -0.0920 | -0.5394 | 0.5023 |

| | | | | | | | | | | | | | |
|---|---|---|---|---|---|---|---|---|---|---|---|---|---|
| | $r^2$ | 0.0240 | 0.0692 | 0.0014 | 0.0001 | 0.0027 | 0.0085 | 0.0592 | 0.0426 | 0.0755 | 0.0085 | 0.2910 | 0.2523 |
| | p-value (u) | 0.4805 | 0.2251 | 0.8631 | 0.9620 | 0.8131 | 0.6760 | 0.2635 | 0.3445 | 0.2045 | 0.6762 | 0.0079 | 0.0146 |
| | p-value (c) | 0.7953 | 0.6813 | 0.9549 | 0.9674 | 0.9519 | 0.8925 | 0.7027 | 0.7267 | 0.6813 | 0.8925 | 0.1517 | 0.1752 |
| **Fz** | r | 0.1622 | 0.2594 | -0.0374 | 0.2492 | 0.2885 | 0.0464 | -0.0841 | 0.1678 | 0.3169 | 0.5028 | 0.1435 | 0.0194 |
| | $r^2$ | 0.0263 | 0.0673 | 0.0014 | 0.0621 | 0.0832 | 0.0022 | 0.0071 | 0.0282 | 0.1004 | 0.2528 | 0.0206 | 0.0004 |
| | p-value (u) | 0.4595 | 0.2320 | 0.8654 | 0.2515 | 0.1818 | 0.8334 | 0.7028 | 0.4439 | 0.1406 | 0.0145 | 0.5135 | 0.9299 |
| | p-value (c) | 0.7953 | 0.6813 | 0.9549 | 0.6898 | 0.6813 | 0.9525 | 0.8996 | 0.7953 | 0.5624 | 0.1752 | 0.7991 | 0.9674 |
| **Cz** | r | 0.1047 | 0.2582 | -0.2739 | 0.0622 | 0.0718 | 0.1427 | 0.0633 | 0.2539 | 0.1205 | **0.6599** | 0.1803 | 0.0912 |
| | $r^2$ | 0.0110 | 0.0667 | 0.0750 | 0.0039 | 0.0051 | 0.0204 | 0.0040 | 0.0645 | 0.0145 | **0.4355** | 0.0325 | 0.0083 |
| | p-value (u) | 0.6344 | 0.2342 | 0.2060 | 0.7781 | 0.7449 | 0.5161 | 0.7742 | 0.2423 | 0.5839 | **0.0006** | 0.4104 | 0.6790 |
| | p-value (c) | 0.8925 | 0.6813 | 0.6813 | 0.9222 | 0.9052 | 0.7991 | 0.9222 | 0.6841 | 0.8624 | **0.0576** | 0.7577 | 0.8925 |
| **Pz** | r | -0.3614 | 0.1462 | -0.3402 | -0.2270 | -0.0109 | -0.0298 | -0.0961 | 0.1664 | 0.2346 | 0.5024 | -0.0186 | 0.2265 |
| | $r^2$ | 0.1306 | 0.0214 | 0.1158 | 0.0515 | 0.0001 | 0.0009 | 0.0092 | 0.0277 | 0.0551 | 0.2524 | 0.0003 | 0.0513 |
| | p-value (u) | 0.0902 | 0.5056 | 0.1122 | 0.2977 | 0.9618 | 0.8952 | 0.6707 | 0.4593 | 0.2812 | 0.0146 | 0.9328 | 0.2986 |
| | p-value (c) | 0.4848 | 0.7991 | 0.5386 | 0.7188 | 0.9674 | 0.9674 | 0.8925 | 0.7953 | 0.7188 | 0.1752 | 0.9674 | 0.7188 |
| **Oz** | r | 0.3644 | 0.4408 | 0.0124 | 0.3607 | 0.4038 | 0.3475 | 0.2703 | -0.3307 | 0.3733 | 0.2706 | 0.1575 | -0.2018 |
| | $r^2$ | 0.1328 | 0.1943 | 0.0002 | 0.1301 | 0.1631 | 0.1207 | 0.0730 | 0.1093 | 0.1394 | 0.0732 | 0.0248 | 0.0407 |
| | p-value (u) | 0.0874 | 0.0353 | 0.9551 | 0.0909 | 0.0560 | 0.1042 | 0.2123 | 0.1233 | 0.0793 | 0.2117 | 0.4729 | 0.3557 |
| | p-value (c) | 0.4848 | 0.3389 | 0.9674 | 0.4848 | 0.4848 | 0.5265 | 0.6813 | 0.5624 | 0.4848 | 0.6813 | 0.7953 | 0.7267 |

**Table S3** Correlation between event feature change and clinical score change after TMS treatment for IDS-SR (a) and PCL-5 scores (b). Correlation values calculated using linear regression and reported with Pearson's correlation coefficient (r); Benjamini–Hochberg-corrected (c) and uncorrected (u) p-values reported from the f-test on the model. Bold values trending (corrected value 0.05 < p < 0.08); bold and italicized values significant (corrected p < 0.05).

# a

| **IDS-SR** | | **Delta/Theta** | | | | **Alpha** | | | | **Beta** | | | |
|---|---|---|---|---|---|---|---|---|---|---|---|---|---|
| | | **Number** | Power | Duration | F-span | **Number** | Power | Duration | F-span | **Number** | Power | Duration | F-span |
| **F3** | r | 0.0688 | -0.4507 | 0.3663 | 0.0406 | -0.0949 | -0.1649 | -0.0465 | -0.1828 | -0.0057 | -0.0304 | 0.1236 | -0.3048 |
| | $r^2$ | 0.0047 | 0.2032 | 0.1342 | 0.0016 | 0.0090 | 0.0272 | 0.0022 | 0.0334 | 0.0000 | 0.0009 | 0.0153 | 0.0929 |
| | p-value (u) | 0.7550 | 0.0309 | 0.0856 | 0.8541 | 0.6666 | 0.4522 | 0.8332 | 0.4038 | 0.9795 | 0.8905 | 0.5742 | 0.1573 |
| | p-value (c) | 0.9088 | 0.4332 | 0.4717 | 0.9646 | 0.8820 | 0.7485 | 0.9644 | 0.7151 | 0.9898 | 0.9745 | 0.8404 | 0.5094 |
| **Fp1** | r | 0.2394 | 0.3431 | -0.2250 | 0.1612 | 0.3496 | 0.4403 | 0.0183 | 0.0747 | 0.2951 | 0.4389 | 0.5290 | -0.2907 |
| | $r^2$ | 0.0573 | 0.1177 | 0.0506 | 0.0260 | 0.1222 | 0.1939 | 0.0003 | 0.0056 | 0.0871 | 0.1926 | 0.2798 | 0.0845 |
| | p-value (u) | 0.2712 | 0.1090 | 0.3020 | 0.4625 | 0.1020 | 0.0355 | 0.9339 | 0.7347 | 0.1717 | 0.0361 | 0.0095 | 0.1784 |
| | p-value (c) | 0.6589 | 0.4717 | 0.6589 | 0.7523 | 0.4717 | 0.4332 | 0.9745 | 0.9088 | 0.5094 | 0.4332 | 0.3040 | 0.5094 |
| **Fpz** | r | 0.0266 | 0.2257 | -0.3619 | 0.3690 | 0.2072 | 0.3556 | 0.1265 | -0.0682 | -0.1221 | 0.3786 | ***0.6832*** | -0.2657 |
| | $r^2$ | 0.0007 | 0.0510 | 0.1310 | 0.1362 | 0.0429 | 0.1265 | 0.0160 | 0.0046 | 0.0149 | 0.1434 | ***0.4668*** | 0.0706 |
| | p-value (u) | 0.9041 | 0.3004 | 0.0897 | 0.0831 | 0.3429 | 0.0959 | 0.5651 | 0.7573 | 0.5790 | 0.0748 | ***0.0003*** | 0.2204 |
| | p-value (c) | 0.9745 | 0.6589 | 0.4717 | 0.4717 | 0.6718 | 0.4717 | 0.8404 | 0.9088 | 0.8404 | 0.4717 | ***0.0288*** | 0.5877 |
| **Fp2** | r | 0.2937 | 0.3770 | -0.0761 | 0.0186 | 0.1926 | 0.3991 | 0.3127 | -0.1237 | -0.0154 | 0.4765 | ***0.6591*** | -0.3582 |

| | | | | | | | | | | | | | |
|---|---|---|---|---|---|---|---|---|---|---|---|---|---|
| | **r²** | 0.0862 | 0.1421 | 0.0058 | 0.0003 | 0.0371 | 0.1593 | 0.0978 | 0.0153 | 0.0002 | 0.2271 | ***0.4344*** | 0.1283 |
| | p-value (u) | 0.1738 | 0.0762 | 0.7299 | 0.9329 | 0.3787 | 0.0592 | 0.1463 | 0.5740 | 0.9444 | 0.0215 | ***0.0006*** | 0.0933 |
| | p-value (c) | 0.5094 | 0.4717 | 0.9088 | 0.9745 | 0.7128 | 0.4717 | 0.5094 | 0.8404 | 0.9749 | 0.4128 | ***0.0288*** | 0.4717 |
| **Fz** | r | -0.0631 | -0.0983 | 0.1535 | -0.0114 | -0.2355 | 0.0463 | 0.1100 | -0.2083 | -0.3162 | -0.1197 | -0.1019 | -0.0418 |
| | **r²** | 0.0040 | 0.0097 | 0.0235 | 0.0001 | 0.0555 | 0.0021 | 0.0121 | 0.0434 | 0.1000 | 0.0143 | 0.0104 | 0.0017 |
| | p-value (u) | 0.7749 | 0.6555 | 0.4845 | 0.9589 | 0.2794 | 0.8338 | 0.6175 | 0.3402 | 0.1415 | 0.5865 | 0.6437 | 0.8500 |
| | p-value (c) | 0.9184 | 0.8820 | 0.7625 | 0.9793 | 0.6589 | 0.9644 | 0.8591 | 0.6718 | 0.5094 | 0.8404 | 0.8820 | 0.9646 |
| **Cz** | r | -0.2456 | -0.2822 | 0.2303 | -0.1884 | -0.3893 | -0.3645 | -0.3105 | -0.2089 | -0.1826 | -0.5025 | -0.2278 | -0.1946 |
| | **r²** | 0.0603 | 0.0796 | 0.0530 | 0.0355 | 0.1516 | 0.1328 | 0.0964 | 0.0436 | 0.0333 | 0.2525 | 0.0519 | 0.0379 |
| | p-value (u) | 0.2586 | 0.1920 | 0.2904 | 0.3894 | 0.0663 | 0.0873 | 0.1493 | 0.3388 | 0.4044 | 0.0145 | 0.2958 | 0.3736 |
| | p-value (c) | 0.6589 | 0.5266 | 0.6589 | 0.7151 | 0.4717 | 0.4717 | 0.5094 | 0.6718 | 0.7151 | 0.3480 | 0.6589 | 0.7128 |
| **Pz** | r | -0.1805 | -0.3113 | 0.3395 | 0.2125 | -0.0006 | 0.0242 | 0.1146 | -0.3036 | -0.3209 | -0.4273 | -0.1742 | 0.0190 |
| | **r²** | 0.0326 | 0.0969 | 0.1153 | 0.0452 | 0.0000 | 0.0006 | 0.0131 | 0.0922 | 0.1030 | 0.1825 | 0.0303 | 0.0004 |
| | p-value (u) | 0.4097 | 0.1482 | 0.1130 | 0.3304 | 0.9978 | 0.9149 | 0.6116 | 0.1696 | 0.1354 | 0.0420 | 0.4267 | 0.9313 |
| | p-value (c) | 0.7151 | 0.5094 | 0.4717 | 0.6718 | 0.9978 | 0.9745 | 0.8591 | 0.5094 | 0.5094 | 0.4480 | 0.7187 | 0.9745 |
| **Oz** | r | -0.1584 | 0.2178 | -0.0937 | 0.0689 | -0.0697 | 0.3417 | 0.0851 | 0.1770 | -0.2894 | -0.0328 | 0.1277 | 0.2312 |
| | **r²** | 0.0251 | 0.0474 | 0.0088 | 0.0047 | 0.0049 | 0.1168 | 0.0072 | 0.0313 | 0.0838 | 0.0011 | 0.0163 | 0.0535 |
| | p-value (u) | 0.4702 | 0.3182 | 0.6707 | 0.7547 | 0.7520 | 0.1105 | 0.6995 | 0.4193 | 0.1804 | 0.8820 | 0.5616 | 0.2884 |
| | p-value (c) | 0.7523 | 0.6718 | 0.8820 | 0.9088 | 0.9088 | 0.4717 | 0.9075 | 0.7187 | 0.5094 | 0.9745 | 0.8404 | 0.6589 |

# b

| **PCL-5** | | **Delta/Theta** | | | | **Alpha** | | | | **Beta** |
|---|---|---|---|---|---|---|---|---|---|---|

| | | **Number** | Power | Duration | F-span | **Number** | Power | Duration | F-span | **Number** | Power | Duration | F-span |
|---|---|---|---|---|---|---|---|---|---|---|---|---|---|
| **F3** | r | 0.1937 | -0.5059 | 0.4618 | 0.0740 | -0.0578 | -0.2412 | 0.1159 | -0.2553 | -0.0196 | -0.0371 | 0.2443 | -0.4112 |
| | **$r^2$** | 0.0375 | 0.2559 | 0.2132 | 0.0055 | 0.0033 | 0.0582 | 0.0134 | 0.0652 | 0.0004 | 0.0014 | 0.0597 | 0.1691 |
| | p-value (u) | 0.3757 | 0.0138 | 0.0265 | 0.7372 | 0.7933 | 0.2676 | 0.5986 | 0.2396 | 0.9291 | 0.8665 | 0.2612 | 0.0513 |
| | p-value (c) | 0.7514 | 0.4176 | 0.4476 | 0.9414 | 0.9414 | 0.6760 | 0.9386 | 0.6760 | 0.9428 | 0.9414 | 0.6760 | 0.4794 |
| **Fp1** | r | 0.2436 | 0.1194 | -0.1116 | 0.3686 | 0.3037 | 0.2962 | -0.0222 | 0.0984 | 0.3065 | 0.3339 | 0.4366 | -0.2708 |
| | **$r^2$** | 0.0593 | 0.0143 | 0.0125 | 0.1359 | 0.0922 | 0.0877 | 0.0005 | 0.0097 | 0.0939 | 0.1115 | 0.1906 | 0.0733 |
| | p-value (u) | 0.2627 | 0.5872 | 0.6122 | 0.0835 | 0.1589 | 0.1700 | 0.9201 | 0.6553 | 0.1550 | 0.1195 | 0.0373 | 0.2114 |
| | p-value (c) | 0.6760 | 0.9386 | 0.9386 | 0.4832 | 0.5671 | 0.5829 | 0.9428 | 0.9386 | 0.5671 | 0.5380 | 0.4476 | 0.6547 |
| **Fpz** | r | -0.0286 | -0.0896 | -0.2281 | 0.3865 | 0.1203 | 0.1833 | 0.0501 | -0.0342 | -0.0919 | 0.2623 | 0.5633 | -0.3033 |
| | **$r^2$** | 0.0008 | 0.0080 | 0.0520 | 0.1494 | 0.0145 | 0.0336 | 0.0025 | 0.0012 | 0.0084 | 0.0688 | 0.3173 | 0.0920 |
| | p-value (u) | 0.8968 | 0.6844 | 0.2952 | 0.0685 | 0.5845 | 0.4025 | 0.8202 | 0.8768 | 0.6767 | 0.2265 | 0.0051 | 0.1595 |
| | p-value (c) | 0.9428 | 0.9386 | 0.6906 | 0.4794 | 0.9386 | 0.7525 | 0.9414 | 0.9414 | 0.9386 | 0.6760 | 0.2448 | 0.5671 |
| **Fp2** | r | 0.3261 | 0.1097 | 0.1871 | 0.0569 | 0.1203 | 0.2092 | 0.2095 | -0.1042 | 0.0377 | 0.3842 | 0.5802 | -0.3654 |
| | **$r^2$** | 0.1063 | 0.0120 | 0.0350 | 0.0032 | 0.0145 | 0.0438 | 0.0439 | 0.0109 | 0.0014 | 0.1476 | 0.3366 | 0.1335 |
| | p-value (u) | 0.1289 | 0.6182 | 0.3927 | 0.7966 | 0.5844 | 0.3381 | 0.3373 | 0.6360 | 0.8643 | 0.0703 | 0.0037 | 0.0864 |
| | p-value (c) | 0.5380 | 0.9386 | 0.7525 | 0.9414 | 0.9386 | 0.6906 | 0.6906 | 0.9386 | 0.9414 | 0.4794 | 0.2448 | 0.4832 |
| **Fz** | r | -0.1221 | -0.4140 | 0.3438 | 0.0461 | -0.2716 | -0.1848 | -0.0125 | -0.2247 | -0.3412 | -0.0959 | -0.0762 | -0.1401 |
| | **$r^2$** | 0.0149 | 0.1714 | 0.1182 | 0.0021 | 0.0737 | 0.0342 | 0.0002 | 0.0505 | 0.1164 | 0.0092 | 0.0058 | 0.0196 |
| | p-value (u) | 0.5787 | 0.0495 | 0.1082 | 0.8344 | 0.2101 | 0.3985 | 0.9549 | 0.3026 | 0.1111 | 0.6632 | 0.7296 | 0.5237 |
| | p-value (c) | 0.9386 | 0.4794 | 0.5333 | 0.9414 | 0.6547 | 0.7525 | 0.9549 | 0.6906 | 0.5333 | 0.9386 | 0.9414 | 0.9141 |
| **Cz** | r | -0.2351 | -0.4422 | 0.3131 | -0.0233 | -0.3267 | -0.3610 | -0.0832 | -0.4524 | -0.2840 | -0.4909 | -0.1021 | -0.2145 |
| | **$r^2$** | 0.0553 | 0.1955 | 0.0981 | 0.0005 | 0.1067 | 0.1303 | 0.0069 | 0.2047 | 0.0806 | 0.2410 | 0.0104 | 0.0460 |

| | | | | | | | | | | | | | |
|---|---|---|---|---|---|---|---|---|---|---|---|---|---|
| | p-value (u) | 0.2802 | 0.0346 | 0.1457 | 0.9158 | 0.1281 | 0.0906 | 0.7060 | 0.0302 | 0.1891 | 0.0174 | 0.6429 | 0.3257 |
| | p-value (c) | 0.6897 | 0.4476 | 0.5671 | 0.9428 | 0.5380 | 0.4832 | 0.9413 | 0.4476 | 0.6260 | 0.4176 | 0.9386 | 0.6906 |
| **Pz** | r | -0.0854 | -0.2152 | 0.3861 | 0.1551 | -0.0412 | -0.0370 | 0.1000 | -0.2340 | -0.2555 | -0.3786 | -0.0943 | -0.0326 |
| | $r^2$ | 0.0073 | 0.0463 | 0.1491 | 0.0241 | 0.0017 | 0.0014 | 0.0100 | 0.0548 | 0.0653 | 0.1433 | 0.0089 | 0.0011 |
| | p-value (u) | 0.6986 | 0.3241 | 0.0688 | 0.4797 | 0.8555 | 0.8702 | 0.6580 | 0.2945 | 0.2394 | 0.0749 | 0.6686 | 0.8826 |
| | p-value (c) | 0.9413 | 0.6906 | 0.4794 | 0.8689 | 0.9414 | 0.9414 | 0.9386 | 0.6906 | 0.6760 | 0.4794 | 0.9386 | 0.9414 |
| **Oz** | r | -0.2471 | -0.0478 | -0.1438 | -0.0513 | -0.2197 | 0.0372 | -0.0354 | 0.0517 | -0.3974 | -0.1813 | 0.0186 | 0.0648 |
| | $r^2$ | 0.0611 | 0.0023 | 0.0207 | 0.0026 | 0.0483 | 0.0014 | 0.0013 | 0.0027 | 0.1579 | 0.0329 | 0.0003 | 0.0042 |
| | p-value (u) | 0.2556 | 0.8287 | 0.5127 | 0.8161 | 0.3137 | 0.8661 | 0.8724 | 0.8148 | 0.0604 | 0.4076 | 0.9330 | 0.7690 |
| | p-value (c) | 0.6760 | 0.9414 | 0.9115 | 0.9414 | 0.6906 | 0.9414 | 0.9414 | 0.9414 | 0.4794 | 0.7525 | 0.9428 | 0.9414 |

**Table S4** Correlation between pre-treatment event feature measurements and clinical score change after TMS treatment for IDS-SR (a) and PCL-5 scores (b). Correlation values calculated using linear regression and reported with Pearson's correlation coefficient (r); Benjamini-Hochberg-corrected (c) and uncorrected (u) p-values reported from the f-test on the model. Bold values trending (corrected value $0.05 < p < 0.08$); bold and italicized values significant (corrected $p < 0.05$).
